# Transportability of missing data models across study sites for research synthesis

**DOI:** 10.64898/2026.03.09.26347913

**Authors:** Robert Thiesmeier, Paul Madley-Dowd, Viktor H. Ahlqvist, Nicola Orsini

## Abstract

**Introduction:** Systematically missing covariates are a common challenge in medical research synthesis of quantitative data, particularly when individual participant data cannot be shared across study sites. Imputing covariate values in studies where they are systematically unobserved using information from sites where the covariate is observed implicitly assumes similarity of associations across studies. The behaviour of this assumption, and the bias arising from violating it, remains difficult to qualitatively reason about. Here, we evaluated a two-stage imputation approach for handling systematically missing covariates using simulations across a range of statistical and causal heterogeneity scenarios.

**Methods:** We conducted a simulation study with varying degrees of between-study heterogeneity and systematic differences in model parameters. A binary confounder was set to systematically missing in half of the studies. Study-specific effect estimates were combined using a two-stage meta-analytic model. The performance of the imputation approach was evaluated with the primary estimand being the pooled conditional confounding-adjusted exposure effect across all studies.

**Results:** Bias in the pooled adjusted effect estimate was small across scenarios with low to substantial between-study heterogeneity. Bias increased monotonically with increasingly pronounced differences in causal structures across study sites. Coverage remained close to the nominal level under low to substantial between-study heterogeneity, but deteriorated markedly as differences in causal structures between study sites became more severe.

**Conclusion:** The two-stage cross-site imputation approach produced valid pooled effect estimates across a wide range of simulated scenarios but showed monotonic sensitivity to differences in causal structures across studies. The results provide insight into the conditions under which cross-site imputation may be appropriate for handling systematically missing covariates in research synthesis.

## 1 Introduction

Collaborative research projects including multiple study sites are increasingly becoming the norm in medical epidemiological research across various domains. The use of individual participant data (IPD), as opposed to synthesis of only aggregated data, allows for estimating confounder adjusted exposure effects from multiple sites, more identifiable diagnostic strategies for individual patients or patient groups, improved identification of risk and prognostic factors, and personalised risk prediction.[15, 14] IPD can be combined across study sites either through collating data into a single dataset or via meta-analysis techniques. In practice, an increasing number of collaborative projects do not share IPD between study sites due to privacy preserving, logistic, or legal barriers.[5] In these circumstances, a two-stage meta-analysis (sometimes also referred to as common data model or federated analysis[5, 8]) can be employed. In a two-stage approach, a common analysis model is fit first at each study site separately. In the second stage, the model estimates and their variances are pooled with a meta-analytical model resulting in a pooled effect estimate across studies. While different names can be used to describe pooling of individual data, we will refer to it as IPD meta-analysis for the remainder of this paper.

Fragmented and non-harmonised data, in particular structurally unrecorded variables (i.e., systematically missing data) across different sites, remain a major challenge in many IPD meta-analyses.[7] Examples include collaborations across international biobanks[10] or electronic health records[3], where certain variables may be systematically unavailable or unmeasured at some sites. Such a problem necessitates either omitting adjustment for key covariates at specific sites or excluding those sites from the analysis altogether. Both approaches are in direct disagreement to the benefits an IPD meta-analysis offers, that is, better possibility for more tailored analysis and/or prediction modelling by using available IPD at all study sites. Additionally, previous studies have shown that ignoring systematically missing data can lead to bias due to lack of confounding adjustment when variables are systematically missing.[21, 17]

Multiple imputation (MI) is one of the most flexible methods to deal with missing data problems. Multilevel MI for hierarchal data structures such as an IPD meta-analysis has been proposed and evaluated to be used for systematically missing data.[1] However, most implementations of current MI strategies require IPD from all studies to be in a single location. Recently, a two-stage MI approach has been proposed to recover variables across sites without the need to pool individual-level data. The two-stage MI approach was outlined in Resche-Rigon and White[12] and further adopted to account for missing data when data across studies cannot be shared.[19] Software is available and described in Thiesmeier, Bottai, Orsini.[20] The approach proceeds by estimating an imputation model in studies with observed covariate data, sharing the resulting regression coefficients and associated variance-covariance structure, combining them via meta-regression, and subsequently imputing the systematically missing covariate in studies without measured data using the imputation variance-covariance structure from the studies with observed data as the basis for imputation. Standard two-stage multivariate meta-analytic methods are then applied to obtain pooled adjusted effect estimates. We will refer to this approach as cross-site imputation hereafter.

Cross-site imputation offers a potential strategy to address a key limitation in most national and international collaborative studies: the inconsistent availability of important covariates across study sites. The validity of this approach relies on a central assumption, namely that the association between the imputation target and the variables used for imputation is sufficiently similar across sites to allow information to be transported between sites. In practice, this transportability assumption may be challenged by between-study differences in populations, measurement processes, or underlying causal structures. Such differences are often considered qualitatively at the design or analysis stage, and their implications for the performance of cross-site imputation are difficult to assess empirically. Therefore, the primary aim of this study is to evaluate the performance of the two-stage cross-site imputation approach by providing evidence from a simulation study that can help in methodological decision making. We consider a range of simulated scenarios with varying degrees of statistical and causal heterogeneity, reflecting plausible departures from the transportability assumption. We thus seek to clarify the circumstances under which cross-site imputation is likely to provide reliable inference in research synthesis settings.

The remainder of the paper is structured as follows. First, the data generating mechanism (DGM), methods, and scenarios used in this simulation study are presented. Second, the results of the simulations are reported. We then introduce and discuss three examples to illustrate cross-site imputation in different scenarios. Last, we discuss the simulation results in the light of the usefulness of the imputation method for systematically missing covariates.

## 2 Methods

### 2.1 Data generating mechanism

Let *S* = 1, …, *K* index *K* studies, with individuals *i* = 1, …, *n*_*s*_ per study. We consider a DGM involving three variables: a continuous exposure *X*_*is*_, a continuous outcome *Y*_*is*_, and a binary confounder *Z*_*is*_ ∈ *{*0, 1*}*. Studies are partitioned into source studies (*S* ∈ *A*) and target studies (*S* ∈ *B*), with *A* ∩ *B* = ∅. In studies *S* ∈ *A*, the confounder *Z*_*is*_ is observed, whereas in studies *S* ∈ *B* the confounder *Z*_*is*_ is systematically missing.

Data for individuals *i* = 1, …, *n*_*s*_ within each study *s* are generated as follows:

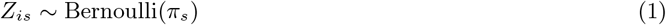

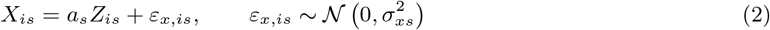

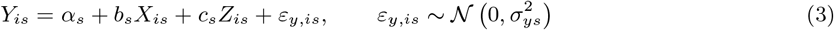

where *σ*_*xs*_ and *σ*_*ys*_ denote study-specific standard deviations of the exposure and outcome. Study-specific parameters vary across studies according to normal distributions centred around hyper-means:

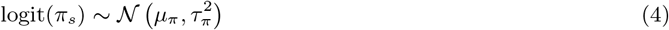

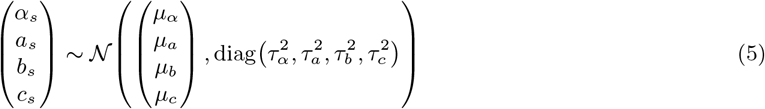

where *µ*_*π*_, *µ*_*α*_, *µ*_*a*_, *µ*_*b*_, *µ*_*c*_ and 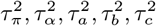 index hyper-means and between-study variances, respectively. Unless otherwise stated, hyper-means are chosen to reflect effect sizes and prevalences commonly encountered in epidemiological and clinical studies. That is, the confounder prevalence is set to 30% (*µ*_*π*_ = logit(0.3)), representing a moderately prevalent binary covariate. Regression coefficients for the exposure-outcome, confounder-outcome, and exposure-confounder associations are set to *µ*_*b*_ = 1, *µ*_*c*_ = 1, and *µ*_*a*_ = 1, respectively. Between-study heterogeneity is parametrised using the standard deviation of the study specific parameters. As such, between-study standard deviations *τ*_*π*_, *τ*_*α*_, *τ*_*a*_, *τ*_*b*_, *τ*_*c*_ are initially set to 0 and varied throughout simulated scenarios (see Table 1). Residual standard deviations of the individual exposure and outcome (*σ*_*xs*_, *σ*_*ys*_) are study-specific and generated as log(*σ*_*xs*_) ∼ 𝒩 0, 0.15^2^, log(*σ*_*ys*_) ∼ 𝒩 0, 0.15^2^, corresponding to moderate variability.

**Table 1:** Overview of the simulated scenarios in this study. Each scenario contains four sub-scenarios. The overview includes a description and definition of how the sub-scenario was implemented in the simulation.

| Scenario |  | Description | Simulation implementation |
| --- | --- | --- | --- |
| **Scenario 1:**<br>Heterogeneity in the parameters of the DGM | Scenario 1.1: Confounder prevalence heterogeneity | Variation in confounder prevalence: $\tau_\pi > 0$ | $\tau_\pi \in \{0, 0.1, 0.2, \dots, 0.6\}$ |
| | Scenario 1.2: Confounding path heterogeneity | Variation in $Z \rightarrow X$ and $Z \rightarrow Y$: $(\tau_a, \tau_c) > 0$ | $(\tau_a, \tau_c) \in \{0, 0.1, 0.2, \dots, 0.6\}$ |
| | Scenario 1.3: Exposure effect heterogeneity | Variation in $X \rightarrow Y$: $\tau_b > 0$ | $\tau_b \in \{0, 0.1, 0.2, \dots, 0.6\}$ |
| | Scenario 1.4: Heterogeneity in all parameters | Variation in all parameters: $(\tau_a, \tau_b, \tau_c, \tau_\alpha, \tau_\pi) > 0$ | $(\tau_a, \tau_b, \tau_c, \tau_\alpha, \tau_\pi) \in \{0, 0.1, 0.2, \dots, 0.6\}$ |
| **Scenario 2:** Systematic shift in the parameters of the DGM between the source and target studies | Scenario 2.1: Shift in prevalence of $Z$ | Different confounder distribution: $\pi_A \neq \pi_B$ | $\pi_B \in \{0.1, 0.2, \dots, 0.6\}$ |
| | Scenario 2.2: Shift in $Z \rightarrow X$ | Different effect of $Z$ on the $X$: $a_A \neq a_B$ | $a_B \in \{0, 0.5, 1, \dots, 3\}$ |
| | Scenario 2.3: Shift in $Z \rightarrow Y$ | Different effect of $Z$ on the $Y$: $c_A \neq c_B$ | $c_B \in \{0, 0.5, 1, \dots, 3\}$ |
| | Scenario 2.4: Shift in both $Z \rightarrow X$ and $Z \rightarrow Y$ | Difference in confounding mechanism: $a_A \neq a_B$, $c_A \neq c_B$ | $(a_B, b_B) \in \{0, 0.5, 1, \dots, 3\}$ |
| **Scenario 3:** Systematic shift and heterogeneity in the parameters of the DGM between the source and target studies | Scenario 3.1: Shift in prevalence of $Z$ + heterogeneity | Different confounder distribution + random variation: $\pi_A \neq \pi_B$, $(\tau_a, \tau_b, \tau_c, \tau_\alpha, \tau_\pi) = 0.3$ | $\pi_B \in \{0, 0.1, 0.2, \dots, 0.6\}$ and $(\tau_a, \tau_b, \tau_c, \tau_\alpha, \tau_\pi) = 0.3$ |
| | Scenario 3.2: Shift in $Z \rightarrow X$ + heterogeneity | Different effect of $Z$ on the $X$ + random variation: $a_A \neq a_B$, $(\tau_a, \tau_b, \tau_c, \tau_\alpha, \tau_\pi) = 0.3$ | $a_B \in \{0, 0.5, 1, \dots, 3\}$ and $(\tau_a, \tau_b, \tau_c, \tau_\alpha, \tau_\pi) = 0.3$ |
| | Scenario 2.3: Shift in $Z \rightarrow Y$ + heterogeneity | Different effect of $Z$ on the $Y$ + random variation: $c_A \neq c_B$, $(\tau_a, \tau_b, \tau_c, \tau_\alpha, \tau_\pi) = 0.3$ | $c_B \in \{0, 0.5, 1, \dots, 3\}$ and $(\tau_a, \tau_b, \tau_c, \tau_\alpha, \tau_\pi) = 0.3$ |
| | Scenario 2.4: Shift in both $Z \rightarrow X$ and $Z \rightarrow Y$ + heterogeneity | Difference in confounding mechanism: $a_A \neq a_B$, $c_A \neq c_B$, $(\tau_a, \tau_b, \tau_c, \tau_\alpha, \tau_\pi) = 0.3$ | $(a_B, b_B) \in \{0, 0.5, 1, \dots, 3\}$ and $(\tau_a, \tau_b, \tau_c, \tau_\alpha, \tau_\pi) = 0.3$ |

After data generation, *Z*_*is*_ was set to systematically missing in half of the studies following a study-level missingness mechanism and commonly used in simulation studies of systematically missing data to generate missing values of such kind.[17, 12, 4]

The DGM induces a strong confounding of the exposure-outcome relationship through *Z*_*is*_, while the effect of *X*_*is*_ on *Y*_*is*_ is non-zero. Inference targets the pooled conditional effect of exposure *X* on the outcome *Y*, controlling for confounding by *Z*. Each study has a sample size of *n*_*s*_ = 1, 000. The causal structure is shown in Figure 1.

**Figure 1:**
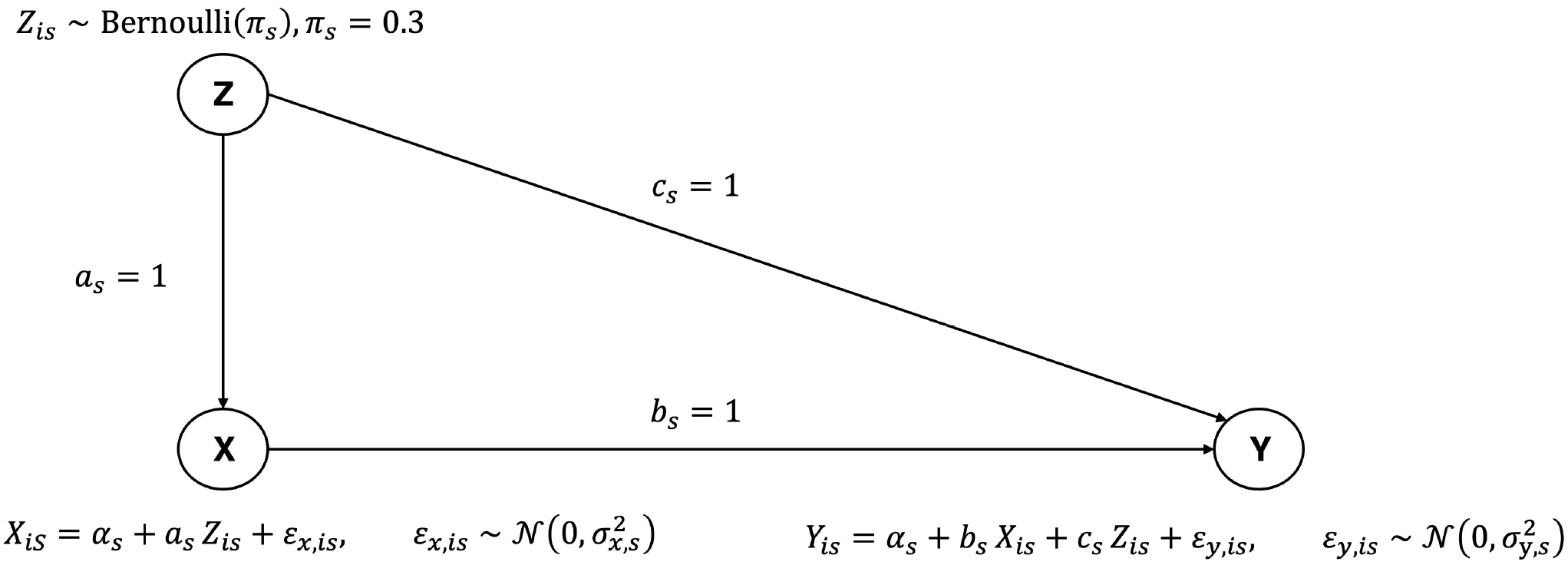
Illustration of the causal structure of the data-generating mechanism where *X, Z*, and *Y* represent the exposure, confounder, and outcome, respectively. The magnitude of the relationship between the parameters is denoted with *a*_*s*_, *b*_*s*_, and *c*_*s*_ where *s* is a study site indicator.

### 2.2 Imputation of systematically missing data

We implemented cross-site imputation as described in Thiesmeier et al.[20] to impute a binary confounder, *Z*_*is*_, which was systematically missing data in target studies (*S* ∈ *B*). In each source study (*S* ∈ *A*) where *Z*_*is*_ is observed, we fit a logistic regression model

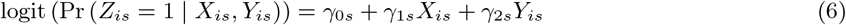

where 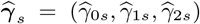 denotes the vector of study-specific regression coefficients with an estimated variance-covariance matrix ***V*** _*s*_.

Next, the study-specific coefficient estimates across source studies are combined using a random-effects meta regression model

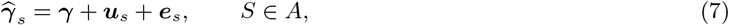

where 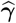 is the pooled coefficient vector. Between-study heterogeneity is modelled via random effects ***u***_*s*_ ∼ 𝒩 (**0, *T***) with

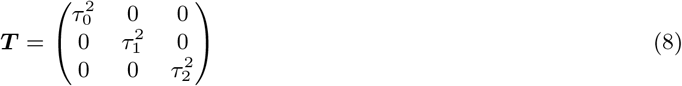

implying independent random effects with coefficient-specific heterogeneity variances. The within-study estimation error is given by ***e***_*s*_ ∼ *N* (**0, *V*** _*s*_). The random effects ***u***_*s*_ and ***e***_*s*_ are assumed independent. Variance components are estimated using restricted maximum likelihood (REML). We denote the pooled estimate by 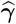 and its variance-covariance matrix 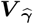.

Importantly, this procedure requires only sharing study-specific estimates 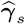 and their variance-covariance matrices ***V*** _*s*_, not requiring sharing IPD between studies.

The last step concludes with the imputation of individual values for the missing covariate *Z*_*is*_ in *S* ∈ *B*. Between-study heterogeneity is incorporated in estimating the pooled coefficients and their uncertainty, while imputation is based on the pooled model. For each imputation *m* = 1, …, *M*, draw a coefficient from the large-sample multivariate normal distribution

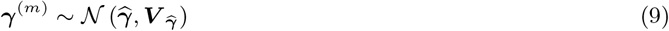

Then, for each individual *i* in target studies *S* ∈ *B*, compute the linear predictor

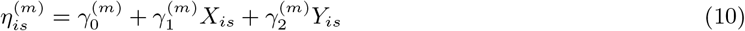

with its corresponding probability

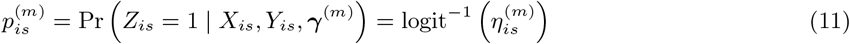

Finally, individual-level imputed values are generated by drawing a random value from a uniform distribution over (0, 1) and assigning 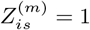 if the draw is less than or equal to 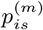, and 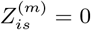 otherwise. Equivalently,

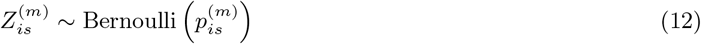

### 2.3 Main analysis

For each simulated dataset, the exposure effect is estimated across all studies using a two-stage univariate random-effects meta-analysis. The following section briefly describes the two stages.

At the first stage, within each study *S* = 1, …, *K*, a linear regression model is fit

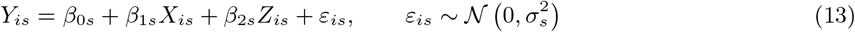

where *Y*_*is*_ denotes the outcome for individual *i* in study *S, X*_*is*_ is the exposure of interest, and *Z*_*is*_ is the confounder adjustment covariate. From each study, the estimated regression coefficients 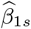 and 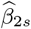, together with their corresponding within-study variances 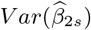 and 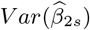 are obtained.

At the second stage, study-specific estimates are pooled separately for each parameter using univariate random-effects meta-analysis models. For *j* ∈ 1, 2,

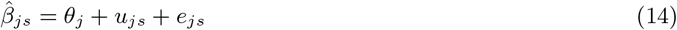

where *θ*_*j*_ denotes the pooled regression coefficient for parameter 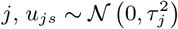 represents between-study heterogeneity, and 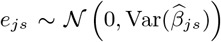 represents within-study sampling error. The between-study variance 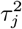 is estimated using REML.

The main parameters are *θ*_1_, and *θ*_2_, corresponding to the pooled conditional exposure effect, and conditional confounder effect, respectively. In particular, *θ*_1_ represents the pooled conditional effect of the exposure on the outcome adjusted for the confounder *Z*, and is typically the primary estimand in research synthesis of observational or randomized studies. The pooled conditional effect of the confounder adjusted for the exposure, *θ*_2_, is primarily assessed for performance evaluation of the imputation-based approach and presented in the Supplementary Materials of this paper.

The data were analysed 1) before data deletion as a benchmark for the MI procedure (reference), 2) after data deletion using complete cases (complete case analysis (CCA)), i.e., restricting the analysis to studies *S* ∈ *A*, 3) in all studies with adjustment *Z*_*is*_ in *S* ∈ *A* but without adjustment for *Z*_*is*_ in *S* ∈ *B* (partial adjustment), and 4) using cross-site imputation.

For analysis method 4), the imputation of *Z*_*is*_ was performed with the `mi impute from` package.[20] After imputation, the analysis model was estimated in each imputed dataset and estimates were combined with Rubin’s rules.[16] For each analysis, *m* = 10 imputations were specified.

The performance of each approach was assessed by computing the empirical mean of the parameter estimates across simulation replications. Bias was defined as the difference between the empirical mean of the parameter estimates and the parameter values specified in the DGM (i.e., 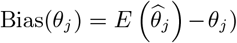. Relative bias (expressed as a percentage, i.e. bias divided by the set parameter value) was also reported to aid interpretability of the results. Additionally, the empirical Monte Carlo standard error (SE) and the coverage of nominal 95% confidence intervals were computed. All analyses were performed with Stata Statistical Software 18 (Stata Corp). The computer code for this simulation study is available in the supplements of this paper.

### 2.4 Transportability assumption of imputation models

Under the presented DGM, transportability refers to the ability to apply an imputation model for the systematically missing confounder *Z* that is learned in source studies (*S* ∈ *A*) to target studies (*S* ∈ *B*). Specifically, we assume that the joint distribution of confounder, exposure, and outcome are identical across source and target studies

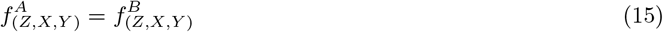

Under this assumption, the imputation model *f*_(*Z*|*X,Y*)_ learned in source studies is correctly specified for target studies. Departures from transportability are introduced by selectively varying parameters of the exposure-confounder and outcome-confounder relationships.

### 2.5 Simulated scenarios

A detailed overview of the simulated scenarios is presented in Table 1. Three categories of scenarios were simulated to assess the performance of cross-site imputation. Parameter values were always changed before missing data was introduced.

#### Scenario 1 Heterogeneity in the parameters of the DGM

We introduce random variation in parameters across all studies without any systematic changes. As such, heterogeneity in individual parameters varied from none to substantial, with (*τ*_*a*_, *τ*_*b*_, *τ*_*c*_, *τ*_*α*_, *τ*_*π*_) ∈ (0, 0.1, 0.2, …, 0.6). Between-study variability is defined relative to the mean parameter values, such that *τ* can be interpreted as a percentage measure of heterogeneity. Accordingly, between-study variability ranged from 0% to 60% of the mean parameter value.

This scenario reflects a common form of heterogeneity encountered in research synthesis, where study-specific parameters vary due to sampling variability, design differences (e.g., study eligibility criteria), or contextual factors, yet are assumed to arise from a common underlying distribution with a shared mean, consistent with the assumptions of random-effects models used in meta-analysis.

#### Scenario 2 Systematic shift in the parameters of the DGM between the source and target studies

We introduce a systematic change in the magnitude of the relationship between parameters of the DGM in target studies (*S* ∈ *B*), but no change in the mean parameter values in source studies (*S* ∈ *A*). As such, the mean parameter values governing the confounder prevalence and the strength of the confounder-exposure and confounder-outcome effects vary systematically between source and target studies (Table 1). Extreme versions of this scenario correspond to either no association in the target studies (mean values of the parameters set to 0) or a three-fold increase in association strength (mean values of the parameters set to 3).

This scenario reflects a more extreme form of heterogeneity in research synthesis, in which the assumption of a shared underlying mean across studies no longer holds. Unlike Scenario 1, these differences are not random fluctuations around a common mean but represent systematic shifts between source and target studies. Such systematic differences may arise from variations in confounding structures due to differences in research settings (e.g., different cultural settings), clinical practices, or measurement processes between study sites.

#### Scenario 3 Systematic shift and heterogeneity in the parameters of the DGM between the source and target studies

In this scenario, we introduce a constant random variation, (*τ*_*a*_, *τ*_*b*_, *τ*_*c*_, *τ*_*α*_, *τ*_*π*_) = 0.30, in parameters across all studies and systematic changes in the magnitude of the relationship between parameters of the DGM in target studies (*S* ∈ *B*). The mean parameter values remain unchanged in source studies (*S* ∈ *A*).

This scenario reflects a hybrid of the previous two settings and imitates a situation in research synthesis where studies display both random variability around study-specific means and systematic differences between study sites. Such situations may arise when combining studies in heterogenous contexts while simultaneously attempting to transport findings across populations with differing confounding structures, clinical practices, or measurement processes.

### 2.6 Heterogeneity between studies

The distinctions, definitions, and implications of differentiating between statistical and causal heterogeneity between the simulated studies requires further clarification. The definition of these two concepts - statistical and causal heterogeneity - for the purposes of this paper thus follows. Here, we define statistical heterogeneity as a change in spread (i.e., standard deviation, *τ*) of the distribution underlying a parameter value. The consequence of such change is an increase in the random variability of the estimates determined in the source and target studies, in addition to an increase in standard error of the pooled effect estimate. Nonetheless, upon repeating the DGM a large number of times, the average values of the distribution within each study remain the same. In contrast, we define causal heterogeneity between studies as a change in the parameter mean (i.e., the effect estimate, *µ*) between the distributions of source and target studies. As such, we introduce a systematic difference between two usets of distributions that does not arise from variability alone. Therefore, upon repeating the DGM a large number of times, we expect to observe a difference in the mean within each study, irrespective of the spread of their distributions.

### 2.7 Sensitivity analyses

We performed three additional sensitivity analyses to assess common modelling assumptions. To facilitate interpretation, all sensitivity analyses were conducted for Scenario 1 only (heterogeneity in the parameters of the DGM).

First, to assess sensitivity to the choice of meta-analytic framework, we replaced the random-effects meta-analysis used in the outcome model with a fixed-effect meta-analysis using inverse-variance weighting. For the final imputation model, we similarly replaced the random-effects specification with a fixed-effect model (i.e., assuming a common coefficient vector across studies), retaining inverse-variance weighting based on the within-study covariance matrices.

Second, to assess sensitivity to the assumption of between-study heterogeneity in the imputation model, we replaced the random-effects imputation model with a fixed-effect meta-regression, while retaining the random-effects univariate meta-analysis for the outcome model.

Third, to evaluate the sensitivity to the number of imputations, we increased the number of imputations to *m* = 50.

In total, we simulated 84 primary scenarios and an additional 84 sensitivity scenarios. Each scenario was simulated 1,000 times.

## 3 Results

In many scenarios, cross-site imputation yielded unbiased estimates of the pooled effect estimate when the systematically missing confounder was imputed using information from source studies. However, the bias increased monotonically, reaching approximately 18%, as systematic differences in the mean values of model parameters between studies became more pronounced. Coverage was close to the nominal level in most scenarios but declined markedly with increasing systematic parameter shifts between source and target sites. Partial adjustment led to consistently larger bias, while CCA lost efficiency compared to the imputation-based approach in many settings.

### 3.1 Scenario 1: Heterogeneity in the parameters of the DGM

We first assessed the performance when substantial variation in parameter values across studies was introduced. Numeric results are presented in Table S1-4.

The bias for the pooled exposure effect (*θ*_1_) after imputing the systematically missing confounder was consistently small across all levels of heterogeneity and stayed below 1% of relative bias (Figure 2). Even under large between-study variation of up to 60% spread in the parameter values, bias for *θ*_1_ after the imputation-based approach and CCA remained negligible and close to that of the reference analysis. Failure to adjust for the confounder in studies where it was systematically missing (partial adjustment) resulted in a larger bias throughout all scenarios (≈8-10%).

**Figure 2:**
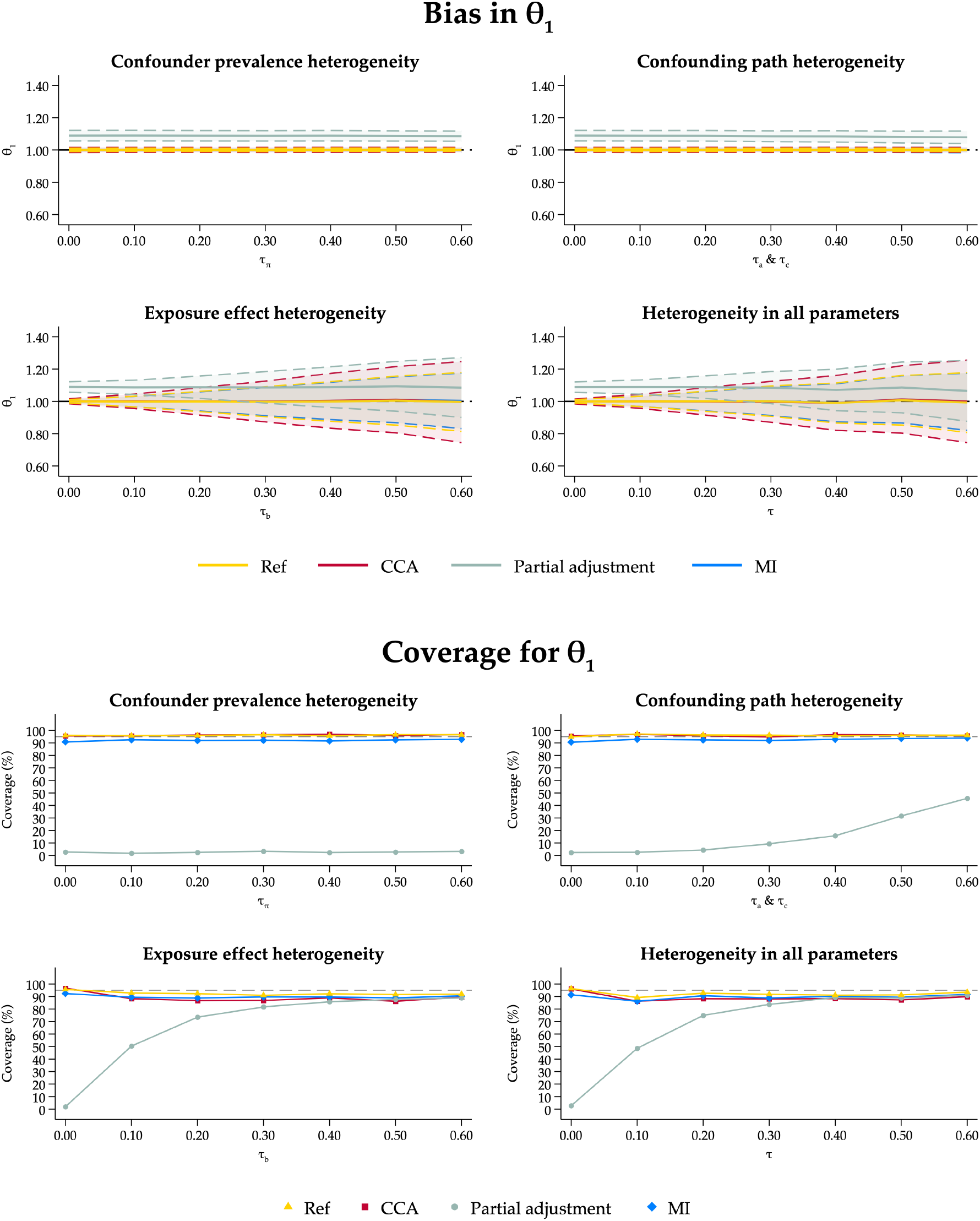
Bias and coverage of 95% confidence intervals for *θ*_1_ in Scenario 2 (Heterogeneity in parameters of the data generating mechanism). Ref: Reference scenario with no missing data; CCA: Complete Case Analysis; MI: Twostage multiple imputation for systemically missing covariates; Partial adjustment: Analysis without adjustment for the systematically missing covariate at target studies.

Under scenarios of large exposure effect heterogeneity (Scenario 1.3) and heterogeneity in all parameters (Scenario 1.4), the empirical SE of 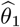 was consistently lower for the imputation-based approach than that of the CCA, suggesting better use of the data. When heterogeneity was very high in all parameters (*τ* = 0.6), the empirical SE for the imputation-based approach was close to the reference (0.190 and 0.189, respectively) and substantially lower than that of the CCA (0.257).

Coverage for the imputation-based approach was slightly higher than that of CCA and close to the reference in all scenarios (Figure 2), being within 5% to the reference values. Even as heterogeneity increased, coverage for *θ*_1_ stayed around 92%, close to the nominal level. Ignoring adjustment for the missing confounder resulted in poorer coverage overall.

The bias was slightly higher for the pooled confounder effect, 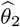, compared to the CCA, however, the magnitude was negligible. The empirical SE for 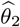 was similar and sometimes slightly larger after MI compared to the CCA (Figure S1). This can be expected given the high fraction of missing data and the resulting inflation of between-imputation variance.

### 3.2 Scenario 2: Systematic shift in the parameters of the DGM between the source and target studies

We next introduced systematic parameter shifts between source and target studies - causal heterogeneity - to examine extreme variation in parameter values that is not due to randomness. The numeric results are presented in Table S5-8.

Across scenarios with systematic shifts in parameter values between source and target studies, bias in the pooled exposure effect (*θ*_1_) were smallest when mean parameter values were close to identical across studies (Figure 3). As systematic differences in parameter values increased, bias in 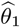 increased monotonically for the imputation-based approach. The bias was high when adjustment for the confounder was ignored in studies with missing data (partial adjustment). Increasing differences in the confounding-path parameters (*a*_*B*_ and *c*_*B*_) led to larger bias in 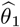 compared to the CCA and ranged from −0.047 (relative bias: ≈5%) to 0.172 (relative bias: ≈17%) when *a*_*B*_ and *c*_*B*_ were set to 0 and 3, respectively, in target studies, compared to 1 in source studies. Despite this large change in the confounding-path parameters, the corresponding increase in relative bias was limited in magnitude.

**Figure 3:**
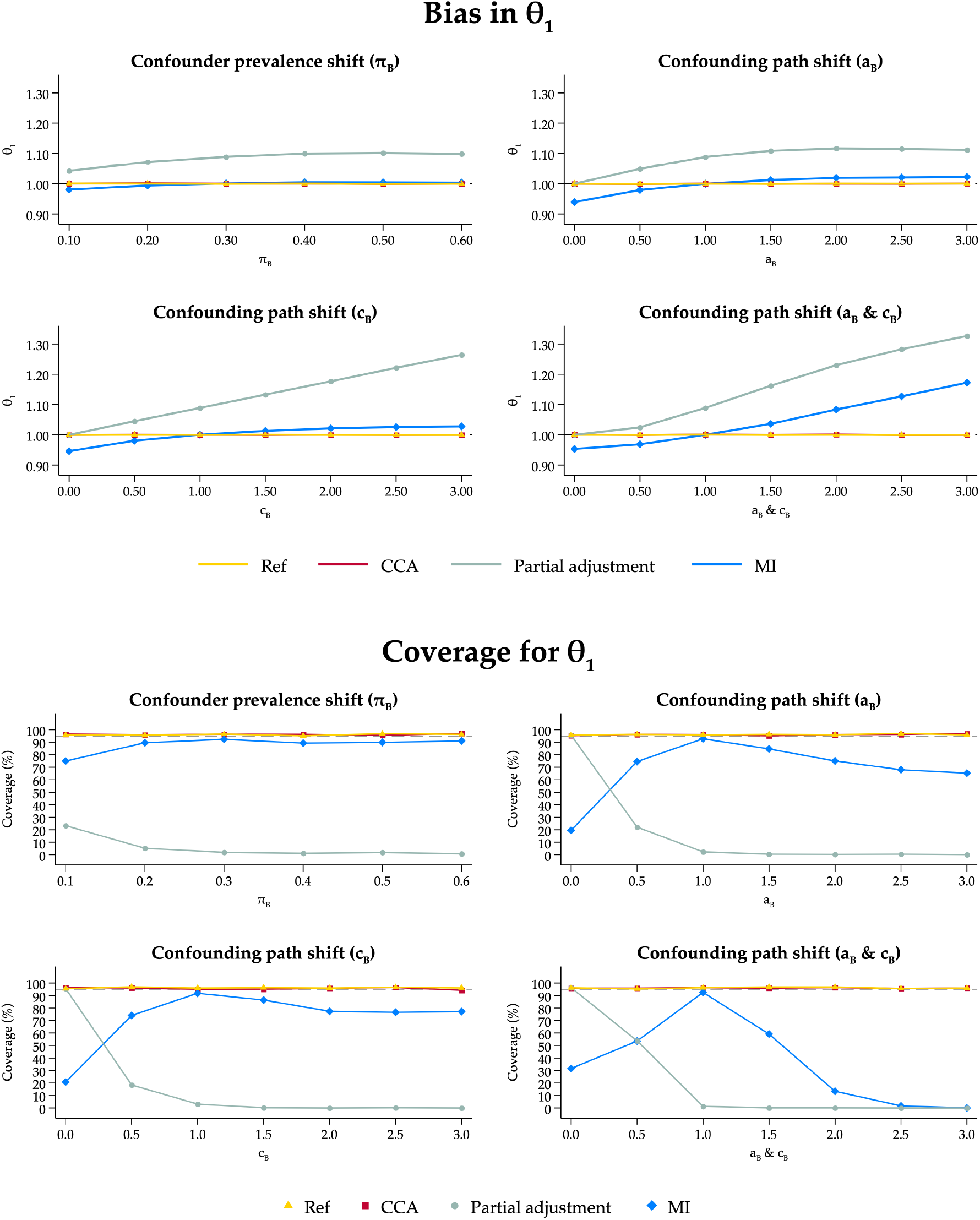
Bias and coverage of 95% confidence intervals for *θ*_1_ and *θ*_2_ in Scenario 2 (Systematic shift in the parameters of the data generating mechanism between the source and target studies). Ref: Reference scenario with no missing data; CCA: Complete Case Analysis; MI: Two-stage multiple imputation for systemically missing covariates; Partial adjustment: Analysis without adjustment for the systematically missing covariate at target studies.

Empirical SEs were comparable between CCA and MI when the magnitude of parameters between source and target studies was similar, however empirical SEs increased for the imputation-based approach when these differences were more pronounced. Coverage for 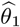 was sensitive to systematic parameter difference. When mean parameter values in target studies were three times larger than those in source studies, reflecting increasing violations of causal similarity across studies, coverage decreased to 0% because of larger bias and inflated empirical SEs (Figure 3). When parameter values were identical across source and target studies, coverage for 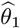 after MI was within 5% to its nominal level.

Similar patterns of bias were observed for 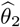, as well as undercoverage (≈82-87%) throughout the scenarios of systematic parameter shifts (Figure S2).

### 3.3 Scenario 3: Systematic shift and heterogeneity in the parameters of the DGM between the source and target studies

Finally, we combined variation in the spread of parameter values and systematic mean parameter shifts to assess the performance in the most complex cases. Between-study heterogeneity in this scenario was moderately high for all parameter values across all studies (*τ* = 0.3), while systematic parameter shifts only occurred at target sites. Numeric results are reported in Table S9-12.

The pattern of bias was similar to the previous scenario, albeit larger variation in the parameters was observed due to the added heterogeneity between studies (Figure 4). Bias in the pooled exposure effect (*θ*_1_) was inflated when systematic shifts in the confounding path parameters (*a*_*B*_ and *c*_*B*_) occurred (ranging from −0.053 (relative bias: ≈5%) when *a*_*B*_ and *c*_*B*_ were set to 0, compared to 0.178 when *a*_*B*_ and *c*_*B*_ were three times larger (relative bias: ≈18%). The bias was highest when no adjustment for the confounder occurred at target sites (up to 32% in relative bias when *a*_*B*_ and *c*_*B*_ were three times larger).

**Figure 4:**
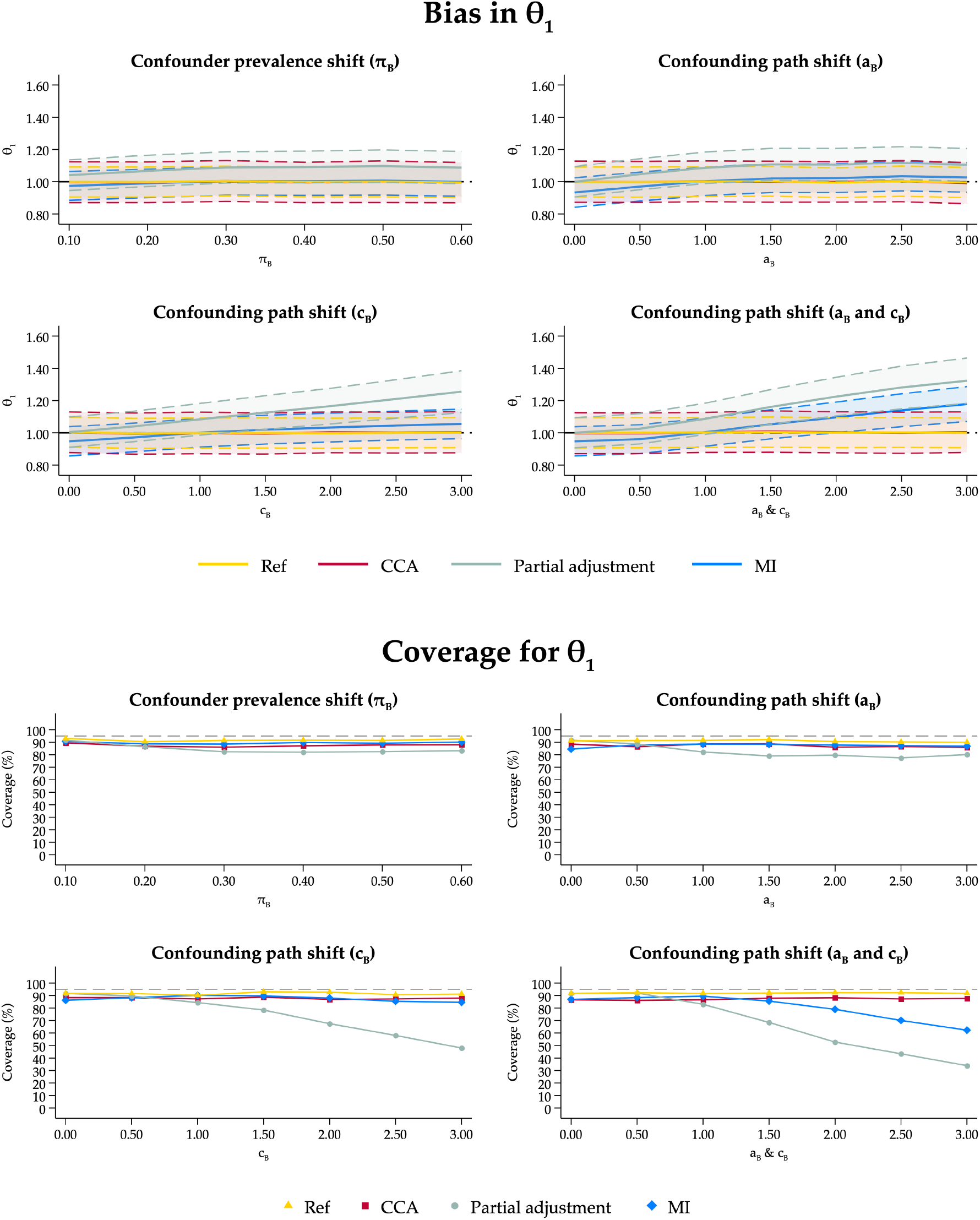
Bias and coverage of 95% confidence intervals in *θ*_1_ in Scenario 3 (Systematic shift and heterogeneity in the parameters of the data generating mechanism between the source and target studies). Ref: Reference scenario with no missing data; CCA: Complete Case Analysis; MI: Two-stage multiple imputation for systemically missing covariates; Partial adjustment: Analysis without adjustment for the systematically missing covariate in target studies.

The empirical SE for the imputation-based approach was lower compared to the CCA, indicating better use of the data in scenarios with low bias. Coverage for 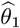 after the imputation-approach was around the nominal value when the parameters did not change systematically between source and target studies (Figure 4). Coverage for the imputation-based approach was slightly higher than the CCA, and within 5% of the reference in all scenarios except when both confounding path parameters differed systematically from the source studies. Here, when *a*_*B*_ and *c*_*B*_ were three times larger, coverage for 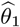 dropped to around 60%. Notably, coverage values in this set of scenarios were higher compared to only introducing systematic changes in the parameter values. This is because the heterogeneity in the parameter values caused additional variation in estimates. This improvement in performance is thus an artifact of noise in the data, masking the systematic parameter shifts between source and target sites.

The bias in the pooled confounder effect (*θ*_2_) remained low. However, coverage for 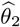 was lower compared to the CCA (Figure S3).

### 3.4 Sensitivity analyses

This section summarises the additional results that were conducted in the sensitivity analyses and are presented in the supplementary materials of this paper.

#### Fixed-effects meta-analytic framework

We applied a fixed-effect meta-analysis with inverse-variance weighting for the outcome model and a fixed-effect meta regression model as the final imputation model. Across all scenarios, bias in 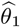 obtained after imputation was consistently higher compared with CCA, however, small in magnitude. Bias for 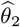 was particularly pronounced when heterogeneity in all parameters of the DGM increased (Figure S4). Coverage was lower for the imputation-based approach relative to the CCA and reference (Figure S5). Notably, coverage decreased rapidly for all approaches as heterogeneity increased, reflecting the lack of flexibility when between-study heterogeneity is not accommodated in either the imputation or outcome model.

#### Fixed-effects meta-regression for the imputation model

We used a fixed-effect meta-regression for pooling imputation-model coefficients, while retaining a univariate random-effects meta-analysis for the outcome model. Bias in 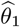 for the imputation-based approach was comparable with other approaches. As heterogeneity increased, bias in 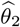 increased more drastically (Figure S6). Coverage for estimates using MI were lower than in the main analysis and declined substantially as heterogeneity increased (Figure S7). The findings highlight that misspecification of the imputation-model heterogeneity primarily affects variance estimation rather than point estimation.

#### Increased number of imputations

This sensitivity analysis mainly aimed at assessing if performance for the imputation-based approach was affected by a higher number of imputations. Bias was not affected and remained small even as heterogeneity increased (Figure S8). A higher number of imputations for the systematically missing confounder did slightly improve the coverage of confidence intervals for the imputation-based approach, being closer to the reference and higher than the CCA (Figure S9). These results suggest that a higher number of imputations can marginally improve some performance measures, however, it comes at an increased computational time.

## 4 Transportability of missing data models between studies: three examples

In the following section, we present three illustrative examples based on simulated data generated from each of the three evaluated scenario categories (Table 1). The examples are intended to demonstrate how patterns commonly interpreted as between-study variability may, under certain DGMs, arise from underlying systematic parameter shifts between source and target studies that violate transportability assumptions. Simulated data are used deliberately, as only under a known DGM is it possible to distinguish heterogeneity in the parameters of the DGM (i.e., differences in the spread of parameter values across studies) from systematic parameter shifts between source and target studies (i.e., systematic differences in mean parameter values across studies). Such distinctions are generally not identifiable in real-world data, where the DGM is unknown. Importantly, these examples are not intended to suggest that systematic parameter shifts between source and target studies can be definitively identified from a single applied dataset. Rather, they illustrate how certain observable features of the data may raise concerns about transportability violations and motivate sensitivity analyses or more cautious interpretation of cross-site imputation results. Data were generated following the same DGM as in the simulations and can be reproduced with the computer code provided in the supplements of this paper.

### 4.1 Example 1: Variability in the spread of the parameters

In the first example, data were generated under substantial between-study heterogeneity in all model parameters (Scenario 1.4, Table 1). Heterogeneity was introduced through random variation in baseline outcome levels, confounder prevalence, and the parameters governing the relationships between exposure, confounder, and outcome.

This example reflects research synthesis based on studies conducted in multiple settings, such as population-based cohort studies from different European countries. While the studies use comparable definitions of exposure, confounders, and outcomes (e.g., registry-based measurements or harmonised questionnaires), they differ in study populations, recruitment strategies, follow-up time, or analytic protocols. As a result, covariate distributions, outcome levels, and effect sizes vary across studies, even though no systematic differences between source and target studies are assumed. Such heterogeneity can for example arise when studies are pooled retrospectively, without having been designed jointly for meta-analysis.

Table 2 summarizes the marginal distributions of the confounder, exposure, and outcome, as well as the crude (unadjusted) exposure effect, for each study and average across studies. Substantial variation is observed across studies in all variables. Nevertheless, all analytical approaches (reference analysis, CCA, partial adjustment, and MI) yielded pooled estimates close to the set effect of *θ*_1_ = 1, with point estimates ranging from 1.11 to 1.30 and corresponding confidence intervals consistently covering the target value (Figure 5, Example 1).

**Table 2:** Descriptive characteristics of the confounder, exposure, and outcome distributions and unadjusted exposure effect estimates across three simulated examples. Descriptive values are presented as percentages for binary and means (SD) for continuous variables across source and target study sites. Data for each example were generated according to the data-generating mechanism described in this paper. Source and target populations each comprised five studies (a total of 10 studies). The pooled unadjusted exposure effect is defined as the effect of the exposure on the outcome without adjustment for an important confounder. In each study, the exposure effect was estimated using a linear regression model, and study-specific estimates and variances were subsequently combined using a univariate random-effects meta-analytic model to obtain pooled estimates across studies. CI, confidence interval; SD, standard deviation

|  |  | Confounder (%) |  | Exposure (mean, SD) |  | Outcome (mean, SD) |  | Unadjusted exposure effect (95% CI) |  |
| --- | --- | --- | --- | --- | --- | --- | --- | --- | --- |
|  |  | Source | Target | Source | Target | Source | Target | Source | Target |
| Example 1 | Study 1 | 41 | 45 | 0.53 (1.47) | 0.22 (1.08) | 1.74 (3.18) | 2.06 (2.27) | 1.96 (1.91, 2.02) | 1.71 (1.63, 1.78) |
|  | Study 2 | 43 | 34 | 0.54 (0.98) | 0.10 (0.91) | 2.40 (2.19) | 1.63 (1.77) | 1.71 (1.62, 1.80) | 1.17 (1.07, 1.27) |
|  | Study 3 | 42 | 20 | 0.28 (1.06) | 0.09 (1.13) | 2.28 (1.88) | 1.22 (1.04) | 1.45 (1.39, 1.52) | 0.58 (0.53, 0.62) |
|  | Study 4 | 42 | 32 | 0.97 (1.38) | 0.28 (1.00) | 2.69 (1.98) | 1.53 (1.87) | 1.28 (1.24, 1.32) | 1.22 (1.13, 1.31) |
|  | Study 5 | 58 | 24 | 0.26 (0.98) | 0.51 (1.52) | 1.97 (1.65) | 1.49 (1.99) | 1.37 (1.31, 1.43) | 0.89 (0.83, 0.95) |
|  | **Average** | **45** | **31** | **0.52 (1.22)** | **0.24 (1.16)** | **2.22 (2.26)** | **1.59 (1.85)** | **1.56 (1.31, 1.80)** | **1.11 (0.74, 1.48)** |
| Example 2 | Study 1 | 31 | 28 | 0.33 (1.08) | 0.84 (1.63) | 1.65 (1.79) | 2.70 (2.98) | 1.18 (1.11, 1.25) | 1.70 (1.66, 1.75) |
|  | Study 2 | 32 | 29 | 0.34 (1.01) | 0.86 (1.65) | 1.65 (1.67) | 2.74 (3.05) | 1.20 (1.13, 1.27) | 1.68 (1.64, 1.73) |
|  | Study 3 | 30 | 31 | 0.29 (1.14) | 0.95 (1.73) | 1.68 (1.76) | 2.91 (3.11) | 1.16 (1.10, 1.22) | 1.59 (1.54, 1.64) |
|  | Study 4 | 29 | 29 | 0.24 (1.15) | 0.81 (1.81) | 1.54 (1.95) | 2.66 (3.16) | 1.23 (1.15, 1.30) | 1.61 (1.57, 1.65) |
|  | Study 5 | 29 | 30 | 0.29 (1.02) | 0.86 (1.69) | 1.60 (1.70) | 2.75 (3.07) | 1.17 (1.10, 1.25) | 1.68 (1.64, 1.72) |
|  | **Average** | **30** | **30** | **0.30 (1.08)** | **0.86 (1.70)** | **1.62 (1.78)** | **2.75 (3.08)** | **1.19 (1.16, 1.22)** | **1.66 (1.61, 1.70)** |
| Example 3 | Study 1 | 38 | 33 | 0.34 (1.02) | 0.75 (1.56) | 1.83 (1.46) | 3.06 (3.10) | 1.01 (0.95, 1.08) | 1.82 (1.77, 1.87) |
|  | Study 2 | 35 | 31 | 0.31 (0.93) | 1.00 (1.76) | 1.62 (1.66) | 2.54 (3.19) | 1.00 (0.91, 1.09) | 1.67 (1.63, 1.72) |
|  | Study 3 | 24 | 26 | 0.07 (0.89) | 0.73 (1.68) | 1.71 (0.98) | 2.63 (2.52) | 0.61 (0.56, 0.67) | 1.30 (1.25, 1.35) |
|  | Study 4 | 19 | 26 | 0.08 (0.94) | 0.88 (1.77) | 1.70 (1.89) | 1.90 (2.90) | 1.61 (1.53, 1.68) | 1.49 (1.45, 1.53) |
|  | Study 5 | 21 | 26 | 0.22 (1.28) | 0.75 (1.70) | 1.54 (1.87) | 2.56 (3.16) | 1.16 (1.11, 1.22) | 1.65 (1.60, 1.70) |
|  | **Average** | **27** | **28** | **0.20 (1.03)** | **0.82 (1.70)** | **1.68 (1.61)** | **2.54 (3.01)** | **1.08 (0.77, 1.39)** | **1.59 (1.41, 1.76)** |

**Figure 5:**
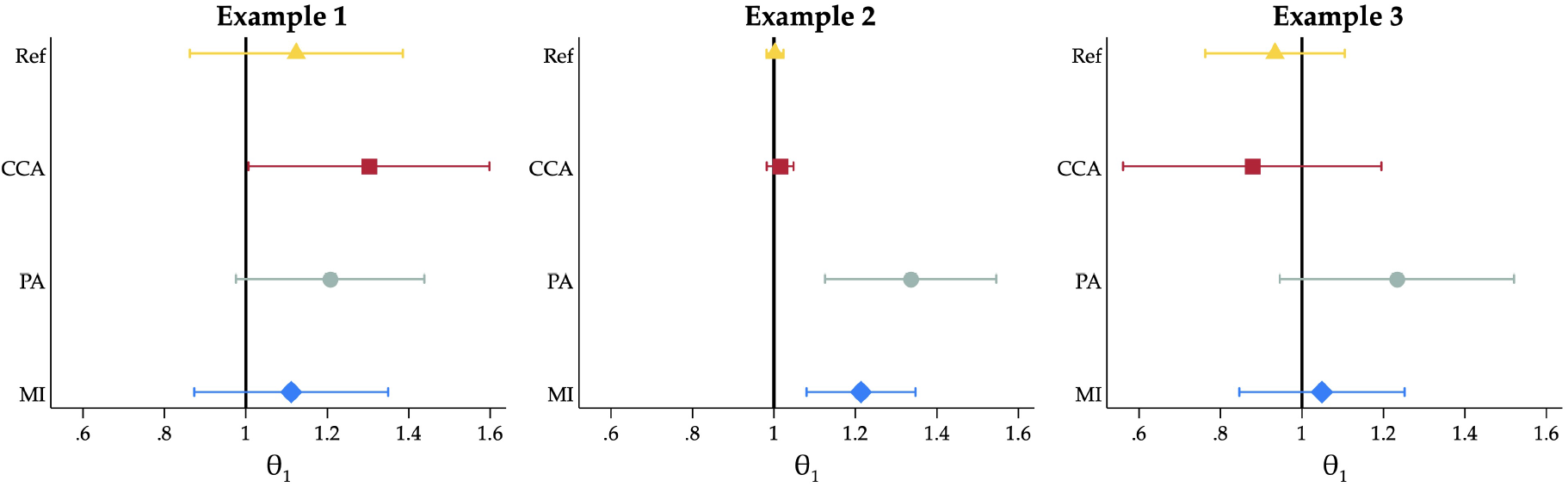
Pooled adjusted estimates for reference (Ref), complete case analysis (CCA), analysis with partial adjustment for the systematically missing covariate (PA), and two-stage multiple imputation for systemically missing covariates (MI). Example 1: Heterogeneity without systematic bias. Example 2: Systematic shifts and induced bias. Example 3: Partial violations. Solid lines indicate the set effect from the DGM.

This example demonstrates that large between-study heterogeneity in model parameters does not in itself induce bias in pooled adjusted estimates, provided that study-specific parameters vary randomly around a common mean (i.e., underlying DGM is shared across studies) and the imputation model is correctly specified.

### 4.2 Example 2: Systematic shifts in the mean of the parameter values

In the second example, data were generated under the conditions of Scenario 2.4 (Table 1). In contrast to Example 1, random heterogeneity was removed and systematic shifts were introduced in parameters governing confounding and outcome generation between source and target studies.

This example reflects a setting with little random variability across studies but pronounced systematic differences between source and target sites. Such situations may arise when studies are pooled across fundamentally different research settings, for example when combining studies conducted in high-income and low- or middle-income countries. In such settings, exposure-confounder and confounder-outcome relationships may differ systematically due to differences in healthcare access, or environmental exposures, even when exposure and outcome definitions are harmonised. For example, the confounding effect of socioeconomic position on both exposure and outcome may be substantially stronger in one setting than another, leading to systematic differences in confounding magnitude that cannot be attributed to random between-study variation.[2]

Table 2 shows that marginal distributions of the confounder, exposure, and outcome were similar within source studies and within target studies, but differed substantially between source and target studies. In particular, the pooled unadjusted exposure effect differed markedly between source and target studies. Because no random between-study heterogeneity in parameter values was introduced in this example, these differences are attributable to systematic shifts in the confounding structure between source and target sites.

In this example, CCA yielded an estimate close to the set effect. In contrast, analyses based on MI and analyses ignoring confounder adjustment in the target studies produced inflated pooled estimates (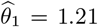 and 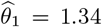, respectively), with narrow confidence intervals excluding the target value (Figure 5, Example 2). Although these results do not provide definitive evidence of non-transportability, the observed patterns are compatible with violations of the transportability assumptions induced by systematic parameter shifts.

### 4.3 Example 3: Systematic shifts in the mean and variability in the spread of the parameter values

In the third example, data were generated under Scenario 3.4 (Table 1), in which systematic parameter shifts between source and target studies were retained while between-study heterogeneity was reintroduced.

This setting extends Example 2 by allowing study-specific parameters to vary randomly within source and target studies, while maintaining systematic differences between the two. Such situations are common in large-scale research synthesis, for example when combining observational studies from multiple countries, where baseline risks, exposure and outcome values, and effect sizes vary across individual studies, as well as confounding structures can differ systematically between regions or healthcare systems. Additional examples include multi-centre studies conducted over long calender periods, where both site-specific variability, temporal changes in clinical practice (e.g., clinical guidelines), or population characteristics (e.g., modified eligibility criteria over time) change.

The marginal distributions of both the outcome and the exposure differed markedly between source and target studies, with limited overlap (Table 2). While such descriptive differences do not constitute definitive evidence of causal differences between study sites, they are consistent with underlying structural differences in the DGMs.

Compared with Example 2, the magnitude of systematic bias is attenuated, reflecting dilution of the structural differences by additional random variation between studies. This is evident from the wider confidence intervals observed for all pooled estimates (Figure 5, Example 3). Bias is not immediately evident from the point estimate, as the estimate obtained after cross-site imputation 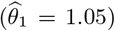 is similar to the reference value 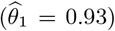, despite underlying differences in confounding structure.

This example illustrates that the performance of cross-site imputation deteriorates gradually rather than abruptly as systematic differences between studies increase and bias is not always immediately evident.

### 4.4 Summary of the examples

Although it is generally impossible to distinguish random variability from systematic shifts in parameter values when the DGM is unknown, several key considerations are highlighted from the examples that mirror the broader results of the simulations. First, distributional differences alone are not diagnostic of bias. As such, differences in confounder, exposure, and outcome distributions can coexist with unbiased estimation when the underlying causal structure is shared (Example 1). Second, bias arises from systematic differences between studies, specifically from shifts in the mean parameter values governing confounding and outcome generation (Example 2). Indications for such systematic difference might be strong differences in effect estimates without adjustment for the systematically missing confounder. Finally, random between-study heterogeneity can partially mask or dilute the impact of systematic differences, leading to more gradual degradation of cross-site imputation performance (Example 3).

## 5 Discussion

There is increasing need for methods to handle missing data in multilevel settings, particularly as IPD meta-analyses and other distributed analyses become more common. In collaborative research, confounder availability often differs across study sites, and IPD may not be shareable. In this paper, we evaluated a two-stage cross-site imputation approach for systematically missing covariate data under a range different heterogeneity scenarios. Overall, the method performed well under low to substantial statistical heterogeneity, including heterogeneity in confounder prevalence and in confounder-exposure/outcome relationships. Performance deteriorated as systematic differences in model parameters between source and target studies increased, most notably through increased bias and reduced confidence-interval coverage. The simulation scenarios were deliberately constructed to include large between-study differences (up to 60% variation in parameter values) to assess the limits of the approach. Even under these conditions, performance was acceptable in many settings.

Our findings are broadly consistent with previous simulation work on multilevel MI. Prior studies reported small bias and mild undercoverage when data were systematically missing[1, 13, 9] and we similarly observed decreasing coverage as heterogeneity in parameter values increased. In scenarios with only systematic parameter shifts between source and target sites, coverage was extremely low. When additional heterogeneity in parameter values was introduced, coverage increased. This pattern is expected because added heterogeneity inflates uncertainty and widens confidence intervals, whereas systematic shifts represent violations of causal similarity that can induce bias without a corresponding increase in estimated uncertainty.

In practice it may be difficult to distinguish statistical heterogeneity from violations of causal similarity as shown in the illustrative examples. In empirical research, confounding structures may differ across study sites. A notable example comes from 1990s birth cohorts examining breastfeeding, where breastfeeding was strongly socially patterned in the UK but less so in Brazil. Socioeconomic position is also associated with BMI, with both the strength and direction of this association varying across contexts.[2] Such heterogeneity in exposure-confounder and confounder-outcome relationships indicates that the confounding structure is not invariant across sites. While this variation can improve causal inference by enabling estimation under distinct confounding regimes, it violates the assumption of structural invariance required for transportability and cross-site imputation.

The results of our simulation study indicate that the cross-site imputation becomes less reliable as systematic parameter differences increase, reflecting reduced transportability across study sites of the missing data models. Related work in distributed data networks evaluated MI approaches when IPD cannot be shared but primarily focused on settings with sporadically missing data[6] and including considerations about the ordering of Rubin’s rules and meta-analysis.[18] Systematically missing covariates have received less attention in these settings. Previous work on systematically missing data in IPD meta-analysis has largely emphasised statistical heterogeneity.[1, 11] Our results highlight that differences in causal structure represent a distinct challenge. In particular, when confounding mechanisms differ across sites (e.g., when confounding path parameters are set to zero in target studies) causal equivalence between sites no longer holds. In such scenarios, negligible bias is expected, however, as we have shown through our simulations, these systematic differences must be very large before materially affecting bias in the pooled exposure effect.

Taken together, these results support the use of cross-site imputation when study sites are expected to be broadly similar in causal structure, even if they differ substantially due to between-study heterogeneity. Conversely, the loss of coverage under increasing systematic parameters shifts between studies underscores the need to consider the plausibility of transportability assumptions before applying the approach. Finally, the trade-off is practical: cross-site imputation often yields smaller bias than ignoring confounding and avoids loss of information relative to CCA but should be applied cautiously when systematic differences between sites are highly plausible.

Some limitations have to be considered. First, although a wide range of scenarios was considered, the DGM necessarily represent simplified versions of real-world settings. More complex structures, such as non-linear effects or effect modification, were not considered. We focused on systematically missing confounders, as this represents a common challenge in research synthesis across multiple study sites.

Second, performance was evaluated primarily within a two-stage meta-analytic framework using a univariate rather than multivariate meta-analysis. Alternative synthesis strategies may behave differently under similar conditions. However, two-stage approaches are required in settings where IPD cannot be shared across sites. As such, the evaluation reflects a realistic and widely used analytical setting. Moreover, a univariate meta-analysis was used to improve numerical stability in settings with a small number of studies and large between-study heterogeneity.

Last, an independent variance-covariance structure was assumed for the random effects in the meta-regression imputation model. In preliminary simulations, models with an unstructured random-effects covariance matrix sporadically resulted in non-negative semidefinite Hessian matrices in scenarios with a limited number of studies, indicating lack of identifiability of variance-covariance parameters. While assuming independence between random effects ignores potential correlations and may lead to underestimated uncertainty for joint inference, we nonetheless used an independent variance-covariance structure to ensure numerical stability and consistent estimation across all simulation scenarios.

## 6 Conclusion

Large collaborative studies that combine evidence across multiple cohorts or trials are increasingly used to address clinical and epidemiological research questions. Such collaborations are often motivated by the need to improve precision of effect estimates, increase statistical power, or evaluate effect heterogeneity across populations. In practice, however, data collection, harmonisation, and variable availability frequently differ across study sites, leading to situations in which key covariates are systematically unmeasured in some studies. At the same time, many research consortia operate under data-sharing constraints that preclude the pooling of IPD. In this setting, methods that can accommodate systematically missing covariates while relying only on summary-level information are essential to enable appropriate confounder adjustment and to make efficient use of all available data. Cross-site imputation provides a principled framework for addressing this challenge. Our simulations have shown that cross-site imputation provides unbiased estimates and makes better use of the data compared to CCA under low to substantial between-study heterogeneity. However, CCA gives less biased estimates when the causal structure differs markedly between source and target studies, leading to more biased estimates after using cross-site imputation. Partial adjustment, that is ignoring adjustment at sites where the confounder is systematically missing, always leads to higher bias and worse performance.

Several directions for future work remain. These include extending the framework to multivariate imputation models, assessing sensitivity to misspecification of the imputation mechanism, and developing diagnostic tools to identify violations of transportability of missing data models. This study provides a starting point for these extensions and highlights key considerations for applied researchers facing systematically missing covariate data in research synthesis.

## Acknowledgments

We would like to thank colleagues and audience members for their constructive feedback and discussions following presentations of this work at several conferences, including the Royal Statistical Society International Conference (2024, 2025) and the Annual Meeting of the International Society of Pharmacoepidemiology (2025).

## Ethical approval

N.A.

## Data Availability

The datasets and computer code to replicate the simulations and the applied examples are provided here.

## Supporting information

Complementary results and results for the sensitivity analyses can be found in the Supplementary Materials for this paper.

## Supplementary Materials

### 1 Scenario 1: Heterogeneity in the parameters of the DGM (base scenario)

#### 1.1 Tables of results

**Table S1:**
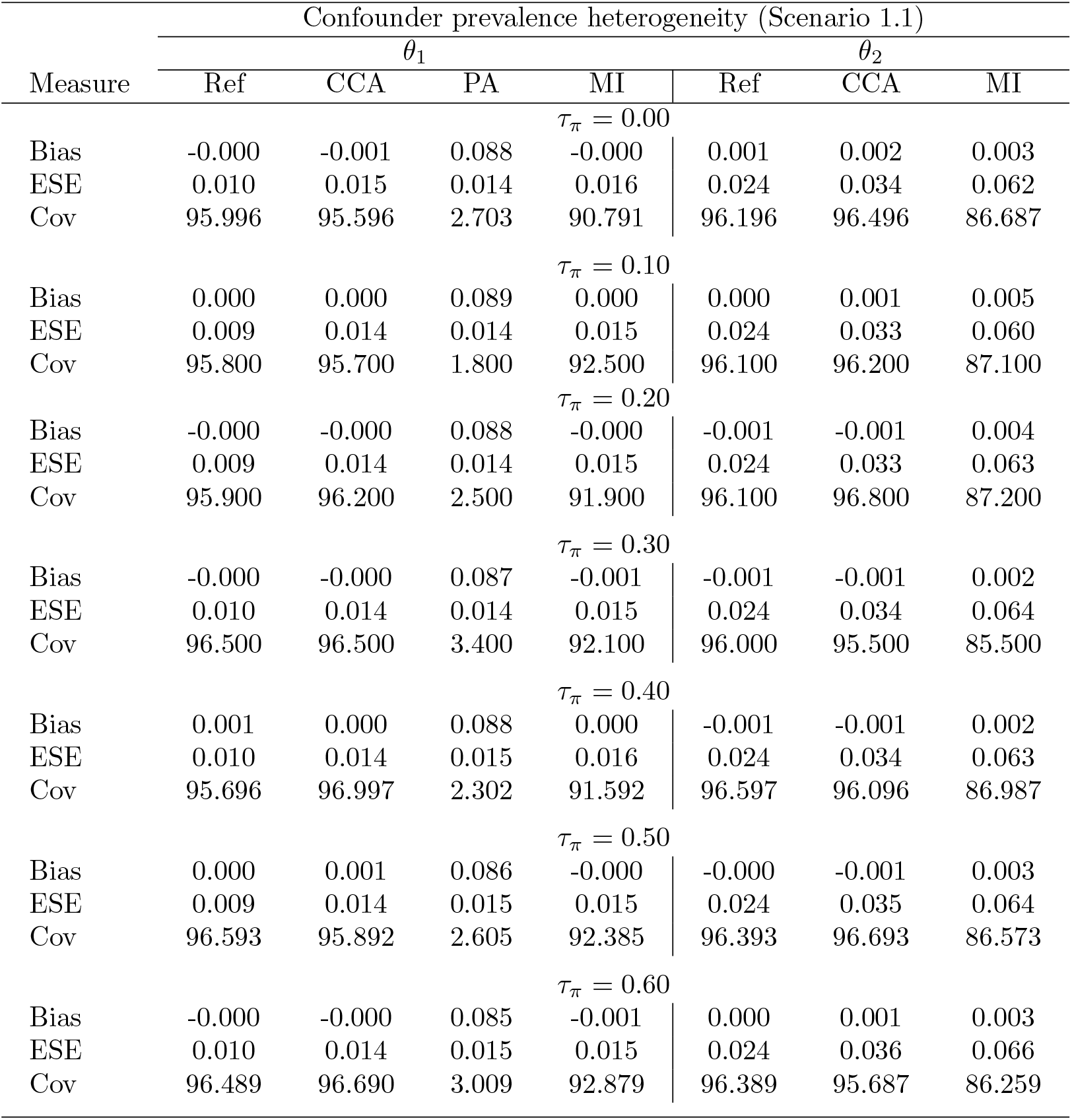
Performance of *θ*_1_ and *θ*_1_ for Scenario 1.1 (prevalence heterogeneity). Ref: Reference scenario with no missing data; CCA: Complete Case Analysis; MI: Two-stage multiple imputation for systemically missing covariates; PA: Analysis without adjustment for the systematically missing covariate in target studies. ESE: Empirical standard error; Cov: Coverage of 95% confidence intervals.

| Measure | Confounder prevalence heterogeneity (Scenario 1.1) |  |  |  |  |  |  |
| --- | --- | --- | --- | --- | --- | --- | --- |
| | $\theta_1$ | | | | $\theta_2$ | | |
|  | Ref | CCA | PA | MI | Ref | CCA | MI |
| $\tau_\pi = 0.00$ | | | | | | | |
| Bias | -0.000 | -0.001 | 0.088 | -0.000 | 0.001 | 0.002 | 0.003 |
| ESE | 0.010 | 0.015 | 0.014 | 0.016 | 0.024 | 0.034 | 0.062 |
| Cov | 95.996 | 95.596 | 2.703 | 90.791 | 96.196 | 96.496 | 86.687 |
| $\tau_\pi = 0.10$ | | | | | | | |
| Bias | 0.000 | 0.000 | 0.089 | 0.000 | 0.000 | 0.001 | 0.005 |
| ESE | 0.009 | 0.014 | 0.014 | 0.015 | 0.024 | 0.033 | 0.060 |
| Cov | 95.800 | 95.700 | 1.800 | 92.500 | 96.100 | 96.200 | 87.100 |
| $\tau_\pi = 0.20$ | | | | | | | |
| Bias | -0.000 | -0.000 | 0.088 | -0.000 | -0.001 | -0.001 | 0.004 |
| ESE | 0.009 | 0.014 | 0.014 | 0.015 | 0.024 | 0.033 | 0.063 |
| Cov | 95.900 | 96.200 | 2.500 | 91.900 | 96.100 | 96.800 | 87.200 |
| $\tau_\pi = 0.30$ | | | | | | | |
| Bias | -0.000 | -0.000 | 0.087 | -0.001 | -0.001 | -0.001 | 0.002 |
| ESE | 0.010 | 0.014 | 0.014 | 0.015 | 0.024 | 0.034 | 0.064 |
| Cov | 96.500 | 96.500 | 3.400 | 92.100 | 96.000 | 95.500 | 85.500 |
| $\tau_\pi = 0.40$ | | | | | | | |
| Bias | 0.001 | 0.000 | 0.088 | 0.000 | -0.001 | -0.001 | 0.002 |
| ESE | 0.010 | 0.014 | 0.015 | 0.016 | 0.024 | 0.034 | 0.063 |
| Cov | 95.696 | 96.997 | 2.302 | 91.592 | 96.597 | 96.096 | 86.987 |
| $\tau_\pi = 0.50$ | | | | | | | |
| Bias | 0.000 | 0.001 | 0.086 | -0.000 | -0.000 | -0.001 | 0.003 |
| ESE | 0.009 | 0.014 | 0.015 | 0.015 | 0.024 | 0.035 | 0.064 |
| Cov | 96.593 | 95.892 | 2.605 | 92.385 | 96.393 | 96.693 | 86.573 |
| $\tau_\pi = 0.60$ | | | | | | | |
| Bias | -0.000 | -0.000 | 0.085 | -0.001 | 0.000 | 0.001 | 0.003 |
| ESE | 0.010 | 0.014 | 0.015 | 0.015 | 0.024 | 0.036 | 0.066 |
| Cov | 96.489 | 96.690 | 3.009 | 92.879 | 96.389 | 95.687 | 86.259 |

**Table S2:** Performance of *θ*_1_ and *θ*_1_ for Scenario 1.2 (confounding path heterogeneity). Ref: Reference scenario with no missing data; CCA: Complete Case Analysis; MI: Two-stage multiple imputation for systemically missing covariates; PA: Analysis without adjustment for the systematically missing covariate in target studies. ESE: Empirical standard error; Cov: Coverage of confidence 95% intervals

| Measure | Confounding path heterogeneity (Scenario 1.2) |  |  |  |  |  |  |
| --- | --- | --- | --- | --- | --- | --- | --- |
| | $\theta_1$ | | | | $\theta_2$ | | |
|  | Ref | CCA | PA | MI | Ref | CCA | MI |
| $\tau_a \& \tau_c = 0.00$ | | | | | | | |
| Bias | 0.000 | 0.000 | 0.089 | 0.001 | -0.001 | -0.000 | 0.001 |
| ESE | 0.010 | 0.014 | 0.014 | 0.016 | 0.024 | 0.034 | 0.063 |
| Cov | 94.895 | 95.495 | 2.302 | 90.591 | 95.996 | 96.597 | 85.185 |
| $\tau_a \& \tau_c = 0.10$ | | | | | | | |
| Bias | -0.001 | -0.001 | 0.088 | -0.000 | 0.000 | 0.000 | 0.007 |
| ESE | 0.009 | 0.013 | 0.014 | 0.015 | 0.040 | 0.057 | 0.076 |
| Cov | 97.189 | 96.787 | 2.209 | 92.871 | 92.470 | 88.655 | 80.723 |
| $\tau_a \& \tau_c = 0.20$ | | | | | | | |
| Bias | 0.000 | 0.000 | 0.087 | 0.001 | -0.003 | -0.004 | 0.002 |
| ESE | 0.010 | 0.014 | 0.017 | 0.016 | 0.066 | 0.093 | 0.101 |
| Cov | 96.289 | 95.787 | 4.112 | 92.377 | 91.876 | 89.468 | 77.834 |
| $\tau_a \& \tau_c = 0.30$ | | | | | | | |
| Bias | 0.000 | -0.000 | 0.084 | -0.000 | -0.001 | 0.005 | 0.011 |
| ESE | 0.010 | 0.014 | 0.019 | 0.017 | 0.097 | 0.137 | 0.134 |
| Cov | 96.092 | 94.790 | 9.218 | 91.884 | 91.283 | 87.475 | 77.355 |
| $\tau_a \& \tau_c = 0.40$ | | | | | | | |
| Bias | 0.000 | 0.001 | 0.084 | 0.001 | 0.004 | 0.004 | 0.015 |
| ESE | 0.010 | 0.013 | 0.024 | 0.017 | 0.127 | 0.178 | 0.169 |
| Cov | 95.696 | 96.597 | 15.716 | 92.893 | 91.892 | 88.488 | 75.475 |
| $\tau_a \& \tau_c = 0.50$ | | | | | | | |
| Bias | 0.000 | -0.000 | 0.080 | -0.000 | -0.003 | 0.003 | 0.010 |
| ESE | 0.009 | 0.013 | 0.027 | 0.018 | 0.159 | 0.227 | 0.214 |
| Cov | 96.092 | 96.293 | 31.463 | 93.487 | 92.585 | 87.275 | 69.639 |
| $\tau_a \& \tau_c = 0.60$ | | | | | | | |
| Bias | -0.001 | -0.000 | 0.078 | -0.001 | -0.004 | -0.010 | 0.003 |
| ESE | 0.010 | 0.014 | 0.032 | 0.019 | 0.196 | 0.272 | 0.257 |
| Cov | 96.073 | 95.770 | 45.317 | 93.857 | 90.836 | 88.822 | 72.407 |

**Table S3:**
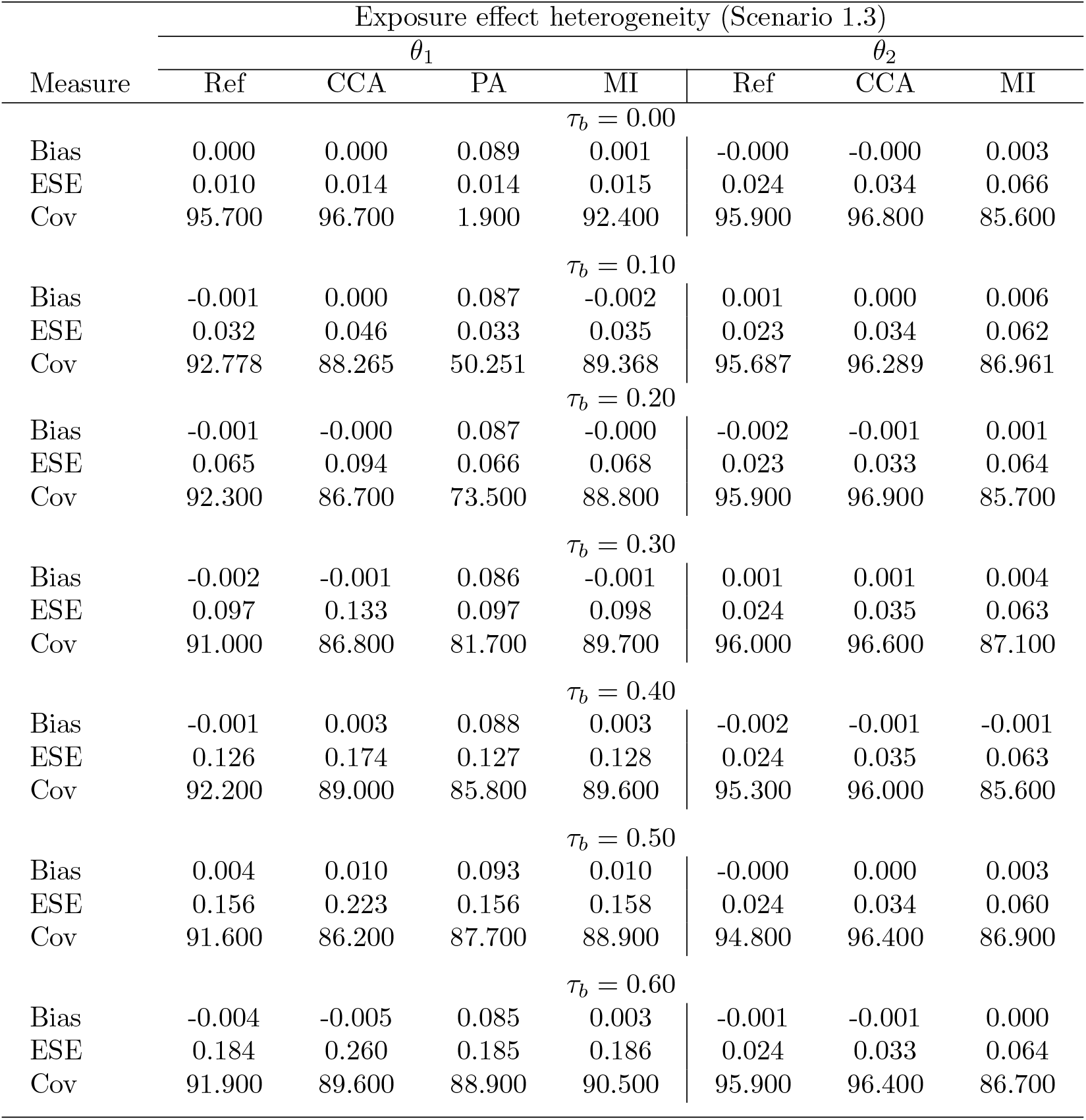
Performance of *θ*_1_ and *θ*_1_ for Scenario 1.3 (effect heterogeneity). Ref: Reference scenario with no missing data; CCA: Complete Case Analysis; MI: Two-stage multiple imputation for systemically missing covariates; PA: Analysis without adjustment for the systematically missing covariate in target studies. ESE: Empirical standard error; Cov: Coverage of 95% confidence intervals.

**Table S4:** Performance of *θ*_1_ and *θ*_1_ for Scenario 1.4 (overall heterogeneity). Ref: Reference scenario with no missing data; CCA: Complete Case Analysis; MI: Two-stage multiple imputation for systemically missing covariates; PA: Analysis without adjustment for the systematically missing covariate in target studies. ESE: Empirical standard error; Cov: Coverage of 95% confidence intervals.

| Measure | Heterogeneity in all parameters (Scenario 1.4) |  |  |  |  |  |  |
| --- | --- | --- | --- | --- | --- | --- | --- |
| | $\theta_1$ | | | | $\theta_2$ | | |
|  | Ref | CCA | PA | MI | Ref | CCA | MI |
| $\tau = 0.00$ | | | | | | | |
| Bias | -0.000 | -0.001 | 0.088 | 0.000 | 0.000 | 0.001 | 0.003 |
| ESE | 0.009 | 0.013 | 0.014 | 0.015 | 0.023 | 0.034 | 0.061 |
| Cov | 96.386 | 96.084 | 2.309 | 91.365 | 95.281 | 96.084 | 87.851 |
| $\tau = 0.10$ | | | | | | | |
| Bias | -0.000 | 0.000 | 0.088 | 0.000 | 0.002 | 0.004 | 0.009 |
| ESE | 0.034 | 0.048 | 0.036 | 0.038 | 0.040 | 0.054 | 0.074 |
| Cov | 89.200 | 86.300 | 48.600 | 86.200 | 92.600 | 91.300 | 84.000 |
| $\tau = 0.20$ | | | | | | | |
| Bias | 0.001 | -0.000 | 0.088 | 0.001 | 0.003 | 0.008 | 0.013 |
| ESE | 0.065 | 0.091 | 0.066 | 0.067 | 0.068 | 0.098 | 0.106 |
| Cov | 92.693 | 88.288 | 74.875 | 90.591 | 90.791 | 87.888 | 78.478 |
| $\tau = 0.30$ | | | | | | | |
| Bias | 0.001 | -0.003 | 0.085 | 0.002 | -0.003 | 0.002 | 0.006 |
| ESE | 0.098 | 0.137 | 0.099 | 0.100 | 0.101 | 0.143 | 0.143 |
| Cov | 91.800 | 88.100 | 83.600 | 88.800 | 89.800 | 86.500 | 75.800 |
| $\tau = 0.40$ | | | | | | | |
| Bias | -0.010 | -0.010 | 0.071 | -0.009 | -0.001 | -0.000 | 0.007 |
| ESE | 0.130 | 0.180 | 0.132 | 0.132 | 0.130 | 0.184 | 0.173 |
| Cov | 91.091 | 88.388 | 89.189 | 90.190 | 91.692 | 87.988 | 75.676 |
| $\tau = 0.50$ | | | | | | | |
| Bias | 0.007 | 0.012 | 0.086 | 0.012 | -0.001 | -0.003 | -0.004 |
| ESE | 0.156 | 0.219 | 0.158 | 0.158 | 0.164 | 0.229 | 0.212 |
| Cov | 91.274 | 87.262 | 88.766 | 89.368 | 91.374 | 88.365 | 72.217 |
| $\tau = 0.60$ | | | | | | | |
| Bias | -0.008 | -0.000 | 0.064 | -0.003 | -0.006 | 0.004 | -0.002 |
| ESE | 0.189 | 0.257 | 0.191 | 0.190 | 0.192 | 0.274 | 0.254 |
| Cov | 93.428 | 89.788 | 90.900 | 91.709 | 91.507 | 88.271 | 71.284 |

#### 1.2 Additional figures

**Figure S1:**
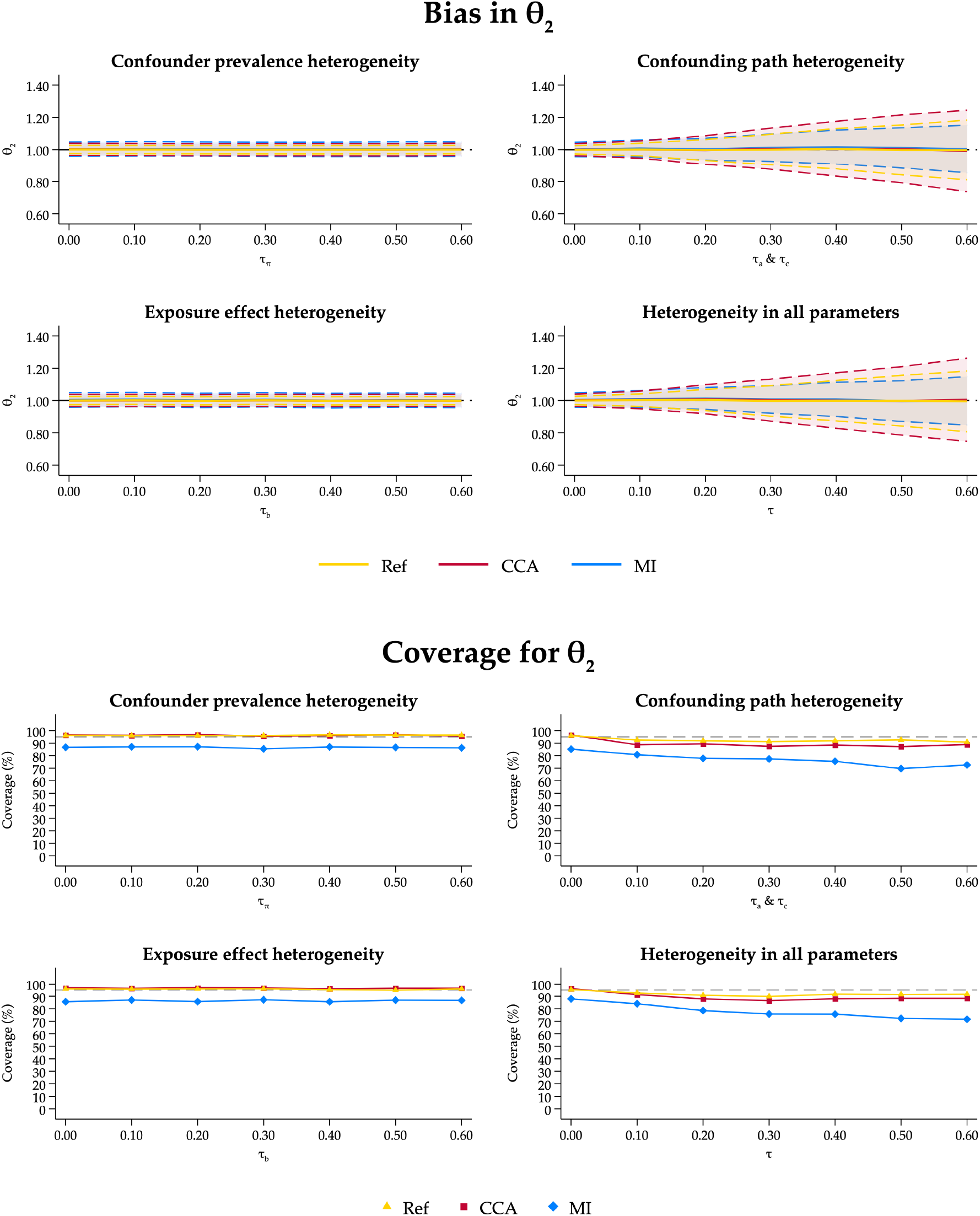
Bias and coverage of 95% confidence intervals for *θ*_2_ in Scenario 1.

### 2 Scenario 2: Systematic shift in the parameters of the DGM between the source and target studies

#### 2.1 Tables of results

**Table S5:**
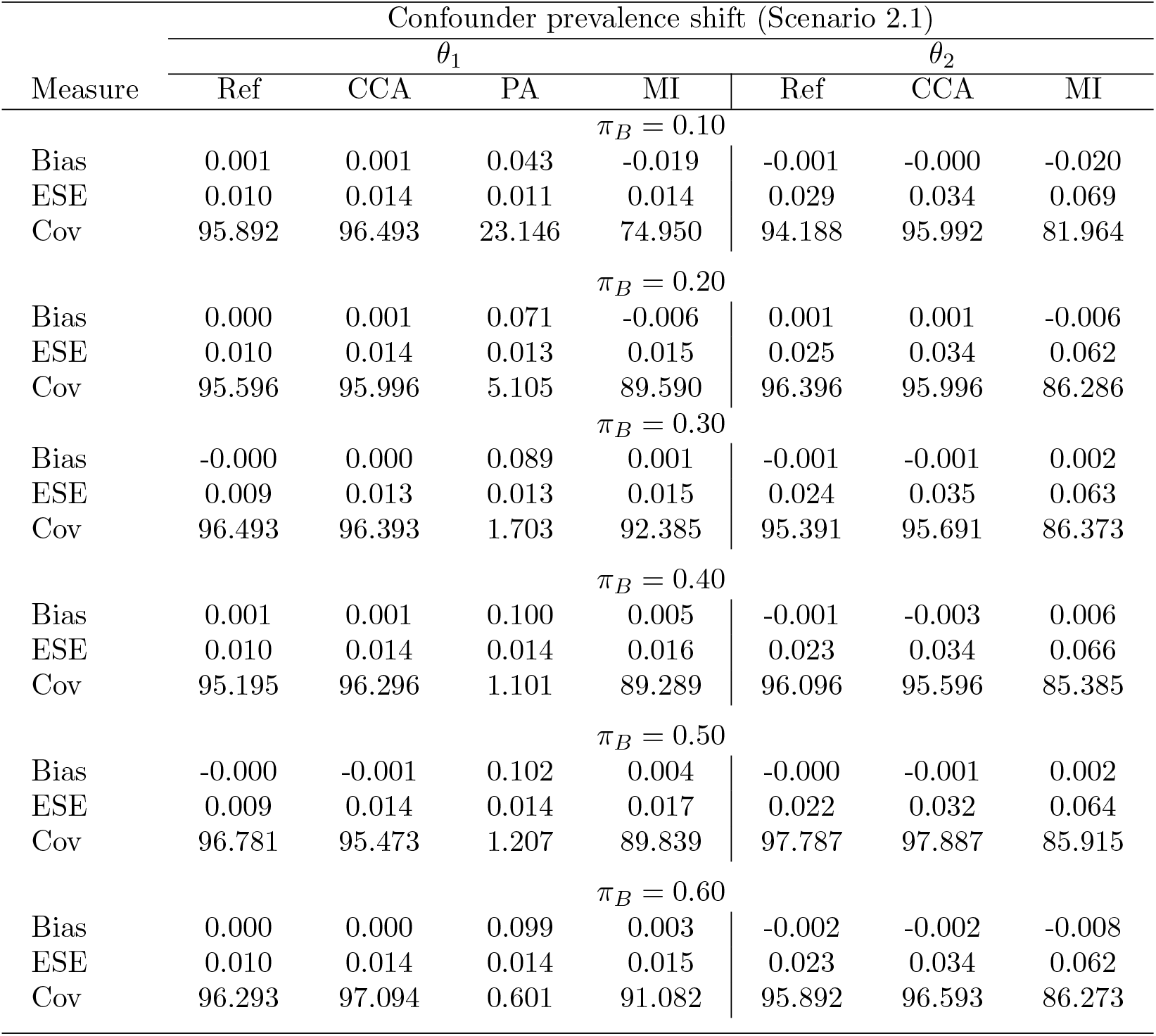
Performance of *θ*_1_ and *θ*_1_ for Scenario 2.1 (prevalence shift). Ref: Reference scenario with no missing data; CCA: Complete Case Analysis; MI: Two-stage multiple imputation for systemically missing covariates; PA: Analysis without adjustment for the systematically missing covariate in target studies. ESE: Empirical standard error; Cov: Coverage of 95% confidence intervals.

**Table S6:** Performance of *θ*_1_ and *θ*_1_ for Scenario 2.2 (prevalence shift). Ref: Reference scenario with no missing data; CCA: Complete Case Analysis; MI: Two-stage multiple imputation for systemically missing covariates; PA: Analysis without adjustment for the systematically missing covariate in target studies. ESE: Empirical standard error; Cov: Coverage of 95% confidence intervals.

| Measure | Confounding path shift ($a_B$) (Scenario 2.2) | | | | | | |
| --- | --- | --- | --- | --- | --- | --- | --- |
| | $\theta_1$ | | | | $\theta_2$ | | |
|  | Ref | CCA | PA | MI | Ref | CCA | MI |
| $a_B = 0.00$ | | | | | | | |
| Bias | -0.000 | -0.000 | 0.000 | -0.061 | 0.000 | 0.000 | 0.014 |
| ESE | 0.010 | 0.014 | 0.010 | 0.017 | 0.022 | 0.033 | 0.065 |
| Cov | 95.800 | 95.300 | 95.700 | 19.500 | 96.700 | 96.700 | 86.100 |
| $a_B = 0.50$ | | | | | | | |
| Bias | -0.001 | -0.001 | 0.049 | -0.021 | -0.001 | -0.002 | 0.005 |
| ESE | 0.010 | 0.014 | 0.012 | 0.016 | 0.024 | 0.034 | 0.064 |
| Cov | 96.300 | 96.300 | 22.100 | 74.600 | 95.000 | 95.800 | 86.400 |
| $a_B = 1.00$ | | | | | | | |
| Bias | 0.000 | 0.001 | 0.088 | 0.000 | -0.000 | -0.000 | 0.005 |
| ESE | 0.010 | 0.014 | 0.013 | 0.015 | 0.023 | 0.032 | 0.062 |
| Cov | 95.884 | 96.084 | 1.908 | 92.771 | 95.884 | 96.888 | 85.944 |
| $a_B = 1.50$ | | | | | | | |
| Bias | -0.000 | -0.000 | 0.109 | 0.012 | 0.000 | -0.000 | 0.000 |
| ESE | 0.009 | 0.014 | 0.013 | 0.017 | 0.024 | 0.033 | 0.063 |
| Cov | 96.293 | 95.190 | 0.301 | 84.669 | 96.693 | 96.593 | 86.874 |
| $a_B = 2.00$ | | | | | | | |
| Bias | 0.001 | 0.000 | 0.117 | 0.019 | -0.000 | 0.000 | -0.006 |
| ESE | 0.010 | 0.014 | 0.012 | 0.017 | 0.027 | 0.034 | 0.061 |
| Cov | 95.992 | 96.092 | 0.100 | 75.050 | 94.489 | 96.393 | 86.072 |
| $a_B = 2.50$ | | | | | | | |
| Bias | 0.000 | -0.000 | 0.115 | 0.021 | -0.003 | -0.001 | -0.010 |
| ESE | 0.010 | 0.014 | 0.011 | 0.017 | 0.027 | 0.033 | 0.062 |
| Cov | 96.991 | 96.289 | 0.201 | 67.904 | 96.690 | 97.292 | 86.760 |
| $a_B = 3.00$ | | | | | | | |
| Bias | 0.000 | 0.001 | 0.112 | 0.022 | -0.000 | -0.001 | -0.015 |
| ESE | 0.009 | 0.014 | 0.010 | 0.015 | 0.028 | 0.033 | 0.061 |
| Cov | 95.896 | 96.897 | 0.000 | 65.265 | 96.096 | 96.496 | 85.085 |

**Table S7:** Performance of *θ*_1_ for Scenario 2.3 (confounding path shift). Ref: Reference scenario with no missing data; CCA: Complete Case Analysis; MI: Two-stage multiple imputation for systemically missing covariates; PA: Analysis without adjustment for the systematically missing covariate in target studies. ESE: Empirical standard error; Cov: Coverage of 95% confidence intervals.

| Measure | Confounding path shift ($c_B$) (Scenario 2.3) | | | |
| --- | --- | --- | --- | --- |
| | $\theta_1$ | | | |
|  | Ref | CCA | PA | MI |
| $c_B = 0.00$ | | | | |
| Bias | -0.000 | -0.000 | -0.000 | -0.054 |
| ESE | 0.010 | 0.014 | 0.010 | 0.015 |
| Cov | 95.500 | 96.400 | 95.500 | 20.800 |
| $c_B = 0.50$ | | | | |
| Bias | 0.000 | 0.000 | 0.045 | -0.019 |
| ESE | 0.010 | 0.014 | 0.011 | 0.015 |
| Cov | 96.891 | 96.088 | 18.255 | 74.022 |
| $c_B = 1.00$ | | | | |
| Bias | -0.000 | -0.000 | 0.089 | 0.000 |
| ESE | 0.010 | 0.014 | 0.013 | 0.015 |
| Cov | 95.996 | 94.995 | 3.003 | 91.792 |
| $c_B = 1.50$ | | | | |
| Bias | 0.000 | -0.000 | 0.133 | 0.013 |
| ESE | 0.010 | 0.014 | 0.018 | 0.018 |
| Cov | 96.200 | 95.100 | 0.200 | 86.400 |
| $c_B = 2.00$ | | | | |
| Bias | -0.000 | 0.000 | 0.177 | 0.021 |
| ESE | 0.010 | 0.014 | 0.022 | 0.022 |
| Cov | 96.000 | 95.900 | 0.000 | 77.400 |
| $c_B = 2.50$ | | | | |
| Bias | -0.000 | -0.000 | 0.221 | 0.026 |
| ESE | 0.009 | 0.014 | 0.026 | 0.026 |
| Cov | 96.593 | 96.493 | 0.000 | 76.553 |
| $c_B = 3.00$ | | | | |
| Bias | -0.000 | -0.000 | 0.264 | 0.028 |
| ESE | 0.010 | 0.015 | 0.031 | 0.028 |
| Cov | 96.100 | 94.200 | 0.000 | 77.200 |

**Table S8:** Performance of *θ*_1_ for Scenario 2.4 (confounding path shift). Ref: Reference scenario with no missing data; CCA: Complete Case Analysis; MI: Two-stage multiple imputation for systemically missing covariates; PA: Analysis without adjustment for the systematically missing covariate in target studies. ESE: Empirical standard error; Cov: Coverage of 95% confidence intervals.

| Measure | Confounding path shift ($a_B$ & $c_B$) (Scenario 2.4) | | | |
| --- | --- | --- | --- | --- |
| | $\theta_1$ | | | |
|  | Ref | CCA | PA | MI |
| $(a_B, c_B) = 0.00$ | | | | |
| Bias | -0.000 | 0.001 | -0.000 | -0.047 |
| ESE | 0.010 | 0.014 | 0.010 | 0.015 |
| Cov | 96.200 | 95.900 | 96.300 | 31.600 |
| $(a_B, c_B) = 0.50$ | | | | |
| Bias | -0.000 | -0.000 | 0.025 | -0.031 |
| ESE | 0.010 | 0.014 | 0.010 | 0.015 |
| Cov | 95.295 | 95.896 | 53.754 | 53.554 |
| $(a_B, c_B) = 1.00$ | | | | |
| Bias | 0.001 | 0.001 | 0.089 | 0.000 |
| ESE | 0.010 | 0.014 | 0.014 | 0.015 |
| Cov | 95.00 | 88.00 | 13.00 | 89.00 |
| $(a_B, c_B) = 1.50$ | | | | |
| Bias | -0.000 | -0.000 | 0.162 | 0.036 |
| ESE | 0.010 | 0.014 | 0.017 | 0.023 |
| Cov | 96.797 | 95.996 | 0.000 | 59.159 |
| $(a_B, c_B) = 2.00$ | | | | |
| Bias | 0.000 | 0.001 | 0.230 | 0.083 |
| ESE | 0.009 | 0.014 | 0.019 | 0.024 |
| Cov | 96.797 | 96.396 | 0.000 | 13.313 |
| $(a_B, c_B) = 2.50$ | | | | |
| Bias | -0.001 | -0.001 | 0.283 | 0.127 |
| ESE | 0.010 | 0.014 | 0.019 | 0.024 |
| Cov | 95.600 | 95.700 | 0.000 | 1.700 |
| $(a_B, c_B) = 3.00$ | | | | |
| Bias | -0.000 | -0.000 | 0.326 | 0.172 |
| ESE | 0.010 | 0.014 | 0.017 | 0.022 |
| Cov | 96.000 | 96.000 | 0.000 | 0.000 |

#### 2.2 Additional figures

**Figure S2:**
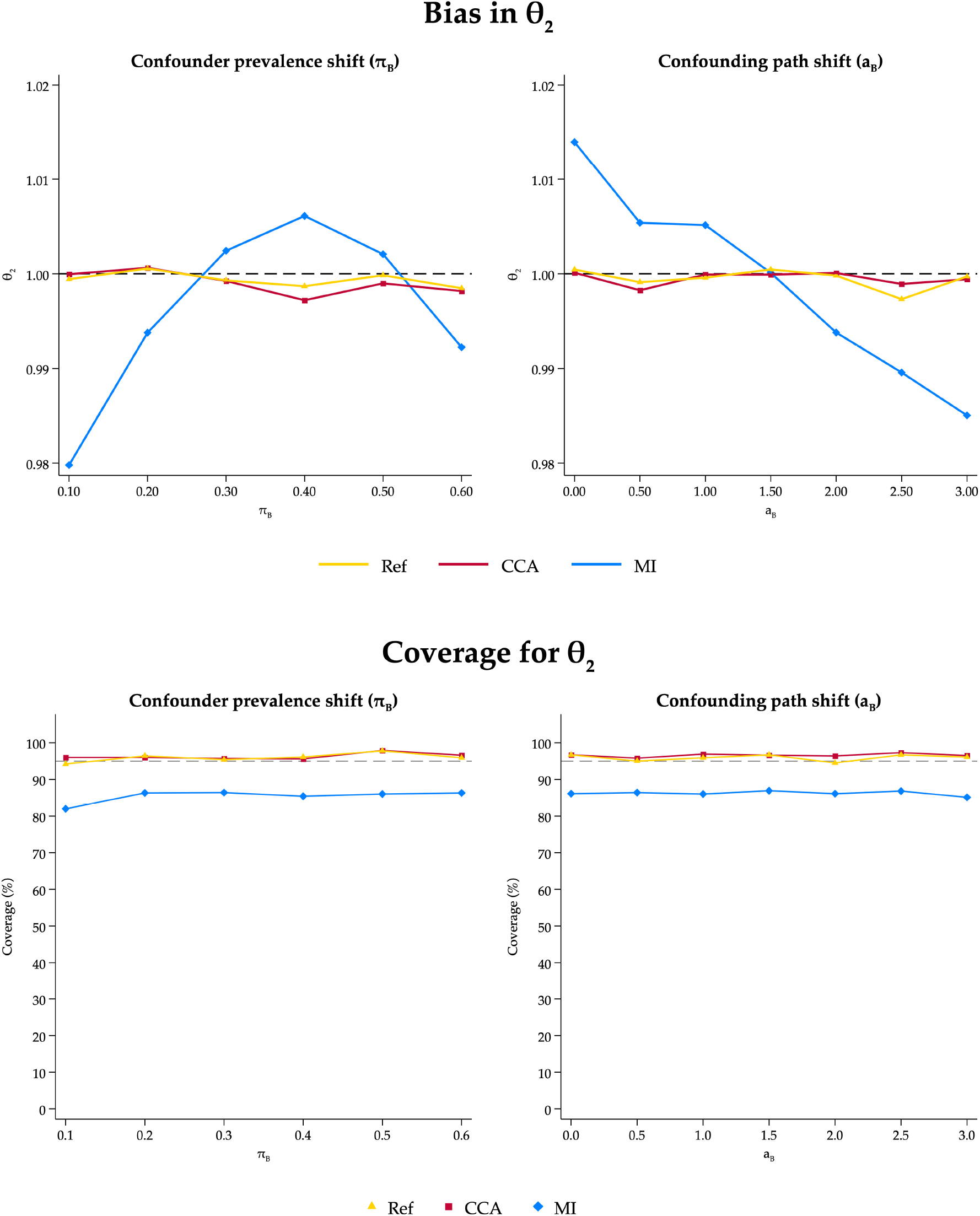
Bias and coverage of 95% confidence intervals for *θ*_2_ in Scenario 2.

### 3 Scenario 3: Systematic shift and heterogeneity in the parameters of the DGM between the source and target studies

#### 3.1 Tables of results

**Table S9:** Performance of *θ*_1_ and *θ*_1_ for Scenario 3.1 (prevalence shift + heterogeneity). Ref: Reference scenario with no missing data; CCA: Complete Case Analysis; MI: Two-stage multiple imputation for systemically missing covariates; PA: Analysis without adjustment for the systematically missing covariate in target studies. ESE: Empirical standard error; Cov: Coverage of 95% confidence intervals.

| Measure | Confounder prevalence shift + heterogeneity (Scenario 3.1) |  |  |  |  |  |  |
| --- | --- | --- | --- | --- | --- | --- | --- |
| | $\theta_1$ | | | | $\theta_2$ | | |
|  | Ref | CCA | PA | MI | Ref | CCA | MI |
| $\pi_B = 0.10$ | | | | | | | |
| Bias | -0.001 | -0.003 | 0.040 | -0.026 | -0.005 | -0.007 | -0.029 |
| ESE | 0.093 | 0.129 | 0.093 | 0.094 | 0.100 | 0.139 | 0.137 |
| Cov | 93.100 | 89.500 | 91.700 | 90.700 | 92.900 | 88.300 | 73.800 |
| $\pi_B = 0.20$ | | | | | | | |
| Bias | -0.002 | -0.004 | 0.067 | -0.010 | 0.004 | -0.001 | 0.000 |
| ESE | 0.097 | 0.142 | 0.098 | 0.100 | 0.098 | 0.140 | 0.136 |
| Cov | 90.500 | 86.900 | 86.400 | 88.700 | 92.000 | 88.800 | 75.200 |
| $\pi_B = 0.30$ | | | | | | | |
| Bias | 0.004 | 0.004 | 0.088 | 0.004 | -0.001 | 0.005 | 0.015 |
| ESE | 0.097 | 0.142 | 0.099 | 0.101 | 0.096 | 0.137 | 0.138 |
| Cov | 91.500 | 86.100 | 82.400 | 88.500 | 92.000 | 88.300 | 75.700 |
| $\pi_B = 0.40$ | | | | | | | |
| Bias | -0.002 | -0.005 | 0.091 | 0.003 | -0.006 | -0.006 | 0.006 |
| ESE | 0.094 | 0.134 | 0.096 | 0.097 | 0.096 | 0.138 | 0.139 |
| Cov | 91.700 | 87.200 | 82.100 | 89.800 | 92.300 | 88.100 | 76.300 |
| $\pi_B = 0.50$ | | | | | | | |
| Bias | 0.000 | -0.000 | 0.096 | 0.007 | -0.000 | -0.002 | 0.007 |
| ESE | 0.097 | 0.133 | 0.097 | 0.098 | 0.099 | 0.141 | 0.139 |
| Cov | 91.500 | 87.900 | 82.500 | 89.200 | 91.500 | 86.600 | 76.200 |
| $\pi_B = 0.60$ | | | | | | | |
| Bias | -0.006 | -0.006 | 0.087 | -0.001 | -0.003 | -0.002 | -0.003 |
| ESE | 0.094 | 0.133 | 0.097 | 0.098 | 0.098 | 0.137 | 0.136 |
| Cov | 92.600 | 88.000 | 83.300 | 90.200 | 92.700 | 87.500 | 76.200 |

**Table S10:** Performance of *θ*_1_ and *θ*_1_ for Scenario 3.2 (confounding path shift + heterogeneity). Ref: Reference scenario with no missing data; CCA: Complete Case Analysis; MI: Two-stage multiple imputation for systemically missing covariates; PA: Analysis without adjustment for the systematically missing covariate in target studies. ESE: Empirical standard error; Cov: Coverage of 95% confidence intervals.

| Measure | Confounding path shift ($a_B$) + heterogeneity (Scenario 3.2) | | | | | | |
| --- | --- | --- | --- | --- | --- | --- | --- |
| | $\theta_1$ | | | | $\theta_2$ | | |
|  | Ref | CCA | PA | MI | Ref | CCA | MI |
| $a_B = 0.00$ | | | | | | | |
| Bias | -0.002 | -0.000 | -0.001 | -0.067 | 0.000 | -0.000 | 0.008 |
| ESE | 0.097 | 0.131 | 0.098 | 0.098 | 0.098 | 0.138 | 0.136 |
| Cov | 91.200 | 88.500 | 91.700 | 84.500 | 92.300 | 88.600 | 77.200 |
| $a_B = 0.50$ | | | | | | | |
| Bias | -0.002 | -0.002 | 0.047 | -0.030 | -0.004 | -0.006 | 0.007 |
| ESE | 0.098 | 0.138 | 0.100 | 0.101 | 0.098 | 0.135 | 0.137 |
| Cov | 91.400 | 86.500 | 88.500 | 87.900 | 91.700 | 88.900 | 77.000 |
| $a_B = 1.00$ | | | | | | | |
| Bias | 0.001 | 0.002 | 0.086 | 0.002 | 0.001 | -0.004 | 0.007 |
| ESE | 0.096 | 0.132 | 0.098 | 0.099 | 0.096 | 0.134 | 0.136 |
| Cov | 91.500 | 88.600 | 82.200 | 88.500 | 92.200 | 88.800 | 76.700 |
| $a_B = 1.50$ | | | | | | | |
| Bias | 0.003 | -0.000 | 0.107 | 0.020 | 0.001 | 0.001 | -0.001 |
| ESE | 0.093 | 0.132 | 0.095 | 0.097 | 0.103 | 0.142 | 0.142 |
| Cov | 92.200 | 88.700 | 79.100 | 88.400 | 90.300 | 86.100 | 72.000 |
| $a_B = 2.00$ | | | | | | | |
| Bias | -0.006 | -0.001 | 0.106 | 0.022 | -0.003 | -0.004 | -0.010 |
| ESE | 0.098 | 0.137 | 0.099 | 0.099 | 0.098 | 0.136 | 0.131 |
| Cov | 90.600 | 86.100 | 79.700 | 87.800 | 90.800 | 87.800 | 78.300 |
| $a_B = 2.50$ | | | | | | | |
| Bias | 0.004 | 0.003 | 0.116 | 0.034 | 0.002 | 0.003 | -0.015 |
| ESE | 0.098 | 0.139 | 0.099 | 0.101 | 0.100 | 0.142 | 0.130 |
| Cov | 90.300 | 86.600 | 77.500 | 87.300 | 91.600 | 86.500 | 76.800 |
| $a_B = 3.00$ | | | | | | | |
| Bias | -0.005 | -0.009 | 0.104 | 0.027 | 0.002 | -0.005 | -0.028 |
| ESE | 0.099 | 0.142 | 0.101 | 0.102 | 0.101 | 0.136 | 0.128 |
| Cov | 90.000 | 85.900 | 80.200 | 86.900 | 91.300 | 89.400 | 77.100 |

**Table S11:** Performance of *θ*_1_ for Scenario 3.3 (confounding path shift + heterogeneity). Ref: Reference scenario with no missing data; CCA: Complete Case Analysis; MI: Two-stage multiple imputation for systemically missing covariates; PA: Analysis without adjustment for the systematically missing covariate in target studies. ESE: Empirical standard error; Cov: Coverage of 95% confidence intervals.

| Measure | Confounding path shift ($c_B$) + heterogeneity (Scenario 3.3) | | | |
| --- | --- | --- | --- | --- |
| | $\theta_1$ | | | |
|  | Ref | CCA | PA | MI |
| | | | $c_B = 0.00$ | |
| Bias | 0.004 | 0.004 | 0.004 | -0.053 |
| ESE | 0.097 | 0.133 | 0.097 | 0.098 |
| Cov | 91.800 | 88.400 | 91.700 | 86.200 |
| | | | $c_B = 0.50$ | |
| Bias | -0.003 | -0.005 | 0.040 | -0.029 |
| ESE | 0.095 | 0.130 | 0.096 | 0.097 |
| Cov | 91.600 | 88.400 | 89.600 | 88.300 |
| | | | $c_B = 1.00$ | |
| Bias | -0.001 | -0.001 | 0.083 | 0.001 |
| ESE | 0.098 | 0.135 | 0.099 | 0.100 |
| Cov | 90.200 | 87.300 | 84.400 | 90.200 |
| | | | $c_B = 1.50$ | |
| Bias | -0.001 | -0.005 | 0.126 | 0.020 |
| ESE | 0.092 | 0.132 | 0.095 | 0.096 |
| Cov | 93.000 | 88.800 | 78.400 | 89.700 |
| | | | $c_B = 2.00$ | |
| Bias | -0.002 | 0.003 | 0.166 | 0.033 |
| ESE | 0.096 | 0.136 | 0.099 | 0.101 |
| Cov | 92.700 | 86.900 | 67.500 | 88.000 |
| | | | $c_B = 2.50$ | |
| Bias | 0.000 | 0.001 | 0.210 | 0.045 |
| ESE | 0.099 | 0.138 | 0.104 | 0.106 |
| Cov | 90.700 | 87.300 | 58.100 | 85.400 |
| | | | $c_B = 3.00$ | |
| Bias | 0.001 | 0.004 | 0.255 | 0.055 |
| ESE | 0.097 | 0.133 | 0.104 | 0.106 |
| Cov | 91.200 | 88.000 | 48.000 | 84.600 |

**Table S12:** Performance of *θ*_1_ for Scenario 3.4 (confounding path shift + heterogeneity). Ref: Reference scenario with no missing data; CCA: Complete Case Analysis; MI: Two-stage multiple imputation for systemically missing covariates; PA: Analysis without adjustment for the systematically missing covariate in target studies. ESE: Empirical standard error; Cov: Coverage of 95% confidence intervals.

| Measure | Confounding path shift ($a_B$ & $c_B$) + heterogeneity (Scenario 3.4) | | | |
| --- | --- | --- | --- | --- |
| | $\theta_1$ | | | |
|  | Ref | CCA | PA | MI |
| | | | $(a_B, c_B) = 0.00$ | |
| Bias | -0.000 | -0.002 | -0.000 | -0.053 |
| ESE | 0.096 | 0.141 | 0.097 | 0.099 |
| Cov | 91.800 | 86.600 | 91.600 | 87.000 |
| | | | $(a_B, c_B) = 0.50$ | |
| Bias | 0.001 | -0.003 | 0.025 | -0.039 |
| ESE | 0.097 | 0.138 | 0.097 | 0.098 |
| Cov | 92.292 | 86.086 | 91.992 | 88.388 |
| | | | $(a_B, c_B) = 1.00$ | |
| Bias | 0.002 | 0.002 | 0.087 | 0.004 |
| ESE | 0.095 | 0.138 | 0.096 | 0.097 |
| Cov | 91.692 | 86.587 | 82.983 | 89.489 |
| | | | $(a_B, c_B) = 1.50$ | |
| Bias | 0.004 | 0.008 | 0.160 | 0.053 |
| ESE | 0.097 | 0.136 | 0.100 | 0.100 |
| Cov | 91.992 | 87.888 | 68.468 | 85.586 |
| | | | $(a_B, c_B) = 2.00$ | |
| Bias | 0.001 | 0.003 | 0.224 | 0.096 |
| ESE | 0.094 | 0.134 | 0.098 | 0.099 |
| Cov | 92.300 | 88.300 | 52.800 | 79.000 |
| | | | $(a_B, c_B) = 2.50$ | |
| Bias | 0.002 | 0.000 | 0.281 | 0.140 |
| ESE | 0.096 | 0.135 | 0.099 | 0.102 |
| Cov | 92.300 | 87.300 | 43.400 | 70.100 |
| | | | $(a_B, c_B) = 3.00$ | |
| Bias | -0.000 | 0.004 | 0.323 | 0.178 |
| ESE | 0.094 | 0.134 | 0.096 | 0.098 |
| Cov | 91.800 | 87.700 | 33.900 | 62.300 |

#### 3.2 Additional figures

**Figure S3:**
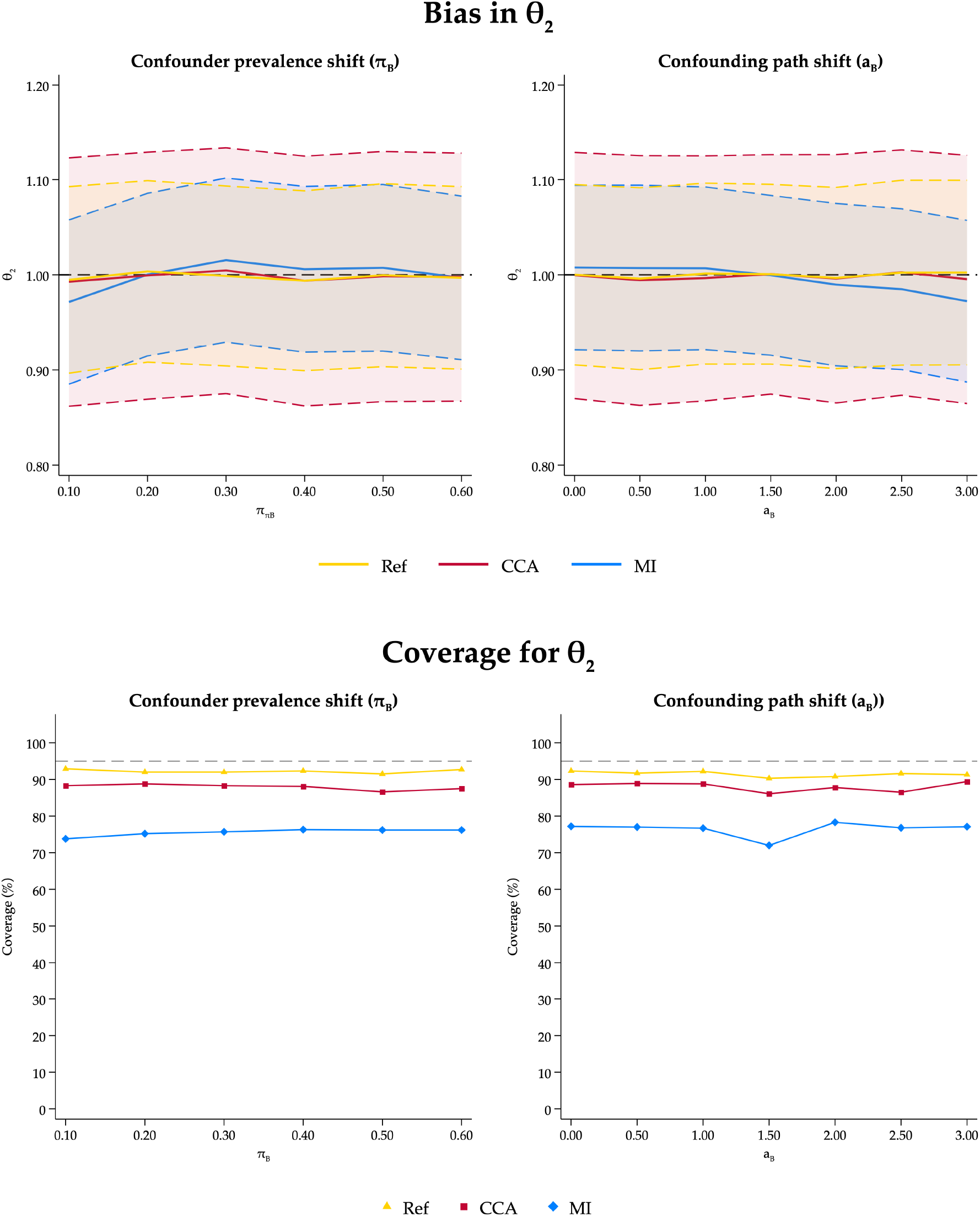
Bias and coverage of 95% confidence intervals for *θ*_2_ in Scenario 3.

### 4 Sensitivity analyses

#### 4.1 Fixed-effects meta-analytic framework

**Figure S4:**
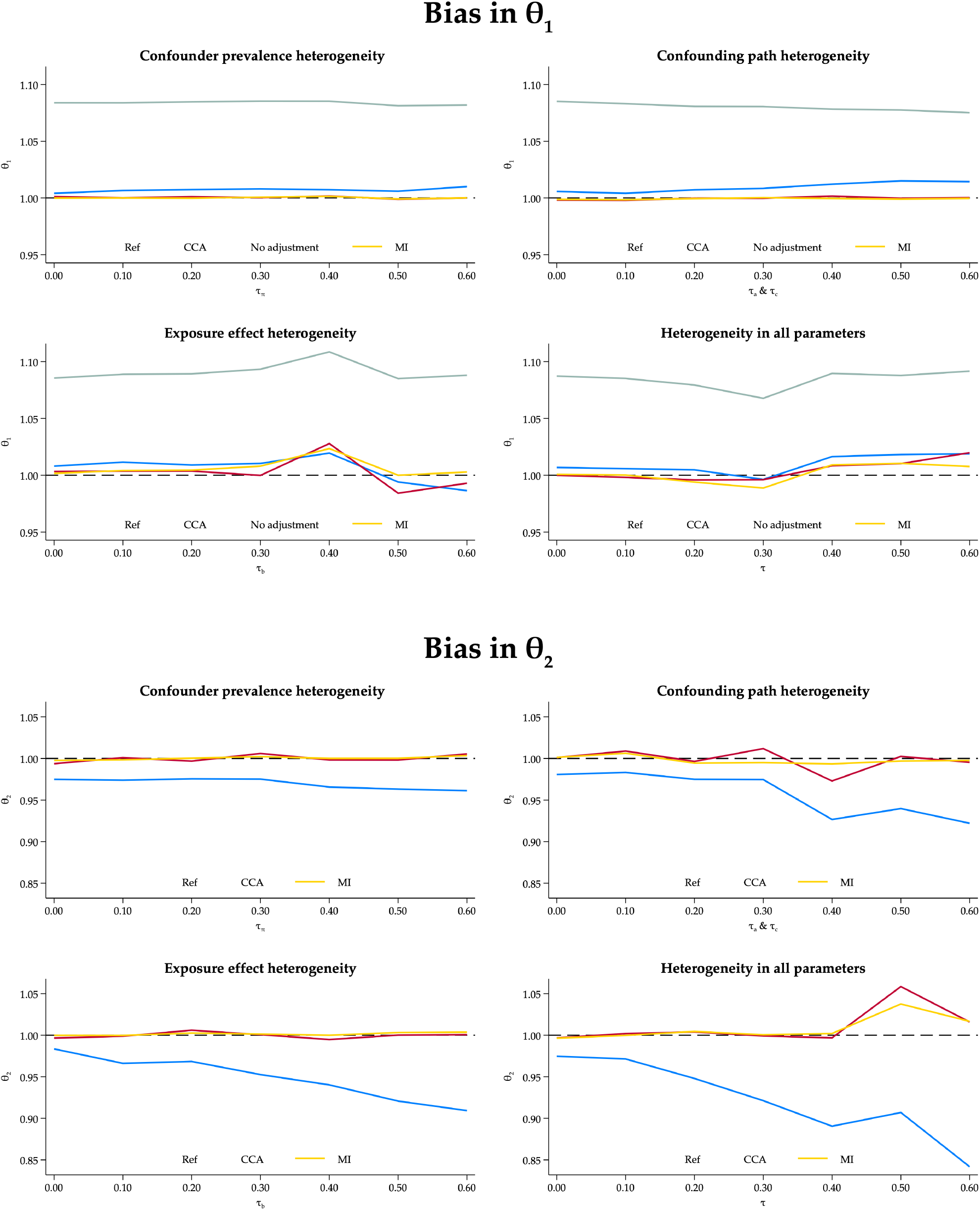
Bias for *θ*_1_ and *θ*_2_

**Figure S5:**
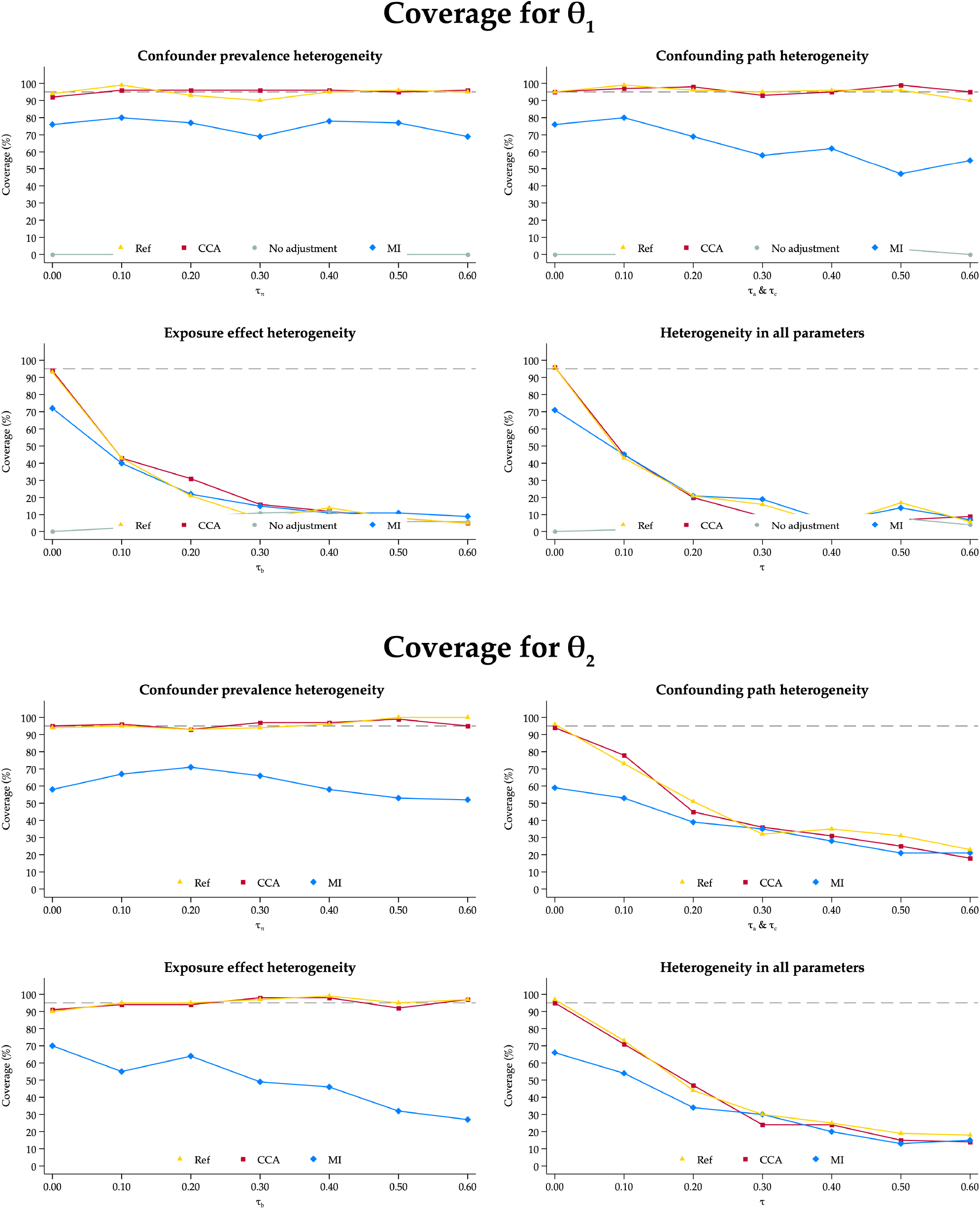
Coverage for *θ*_1_ and *θ*_2_

#### 4.2 Fixed-effects meta-regression for the imputation model

**Figure S6:**
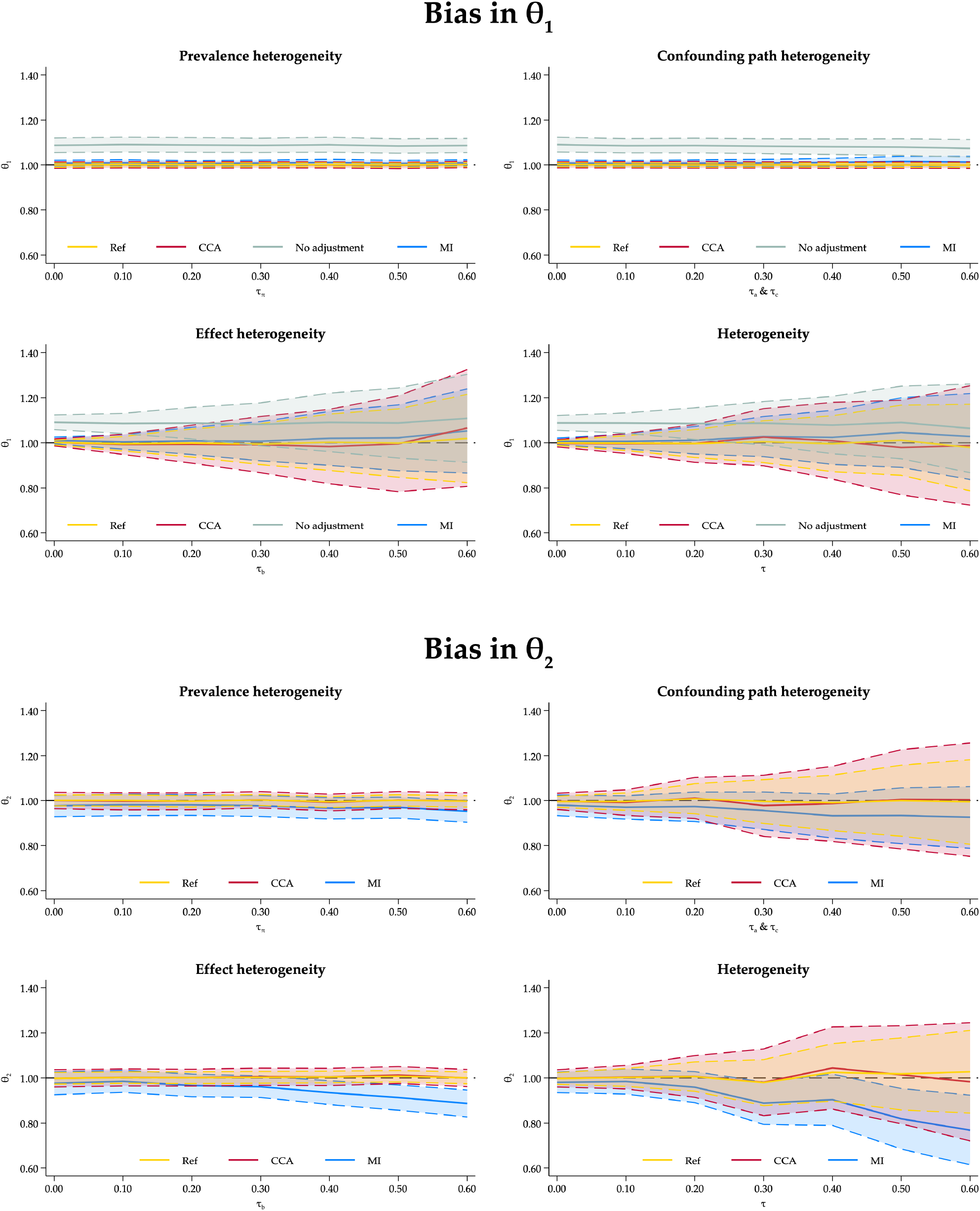
Bias for *θ*_1_ and *θ*_2_

**Figure S7:**
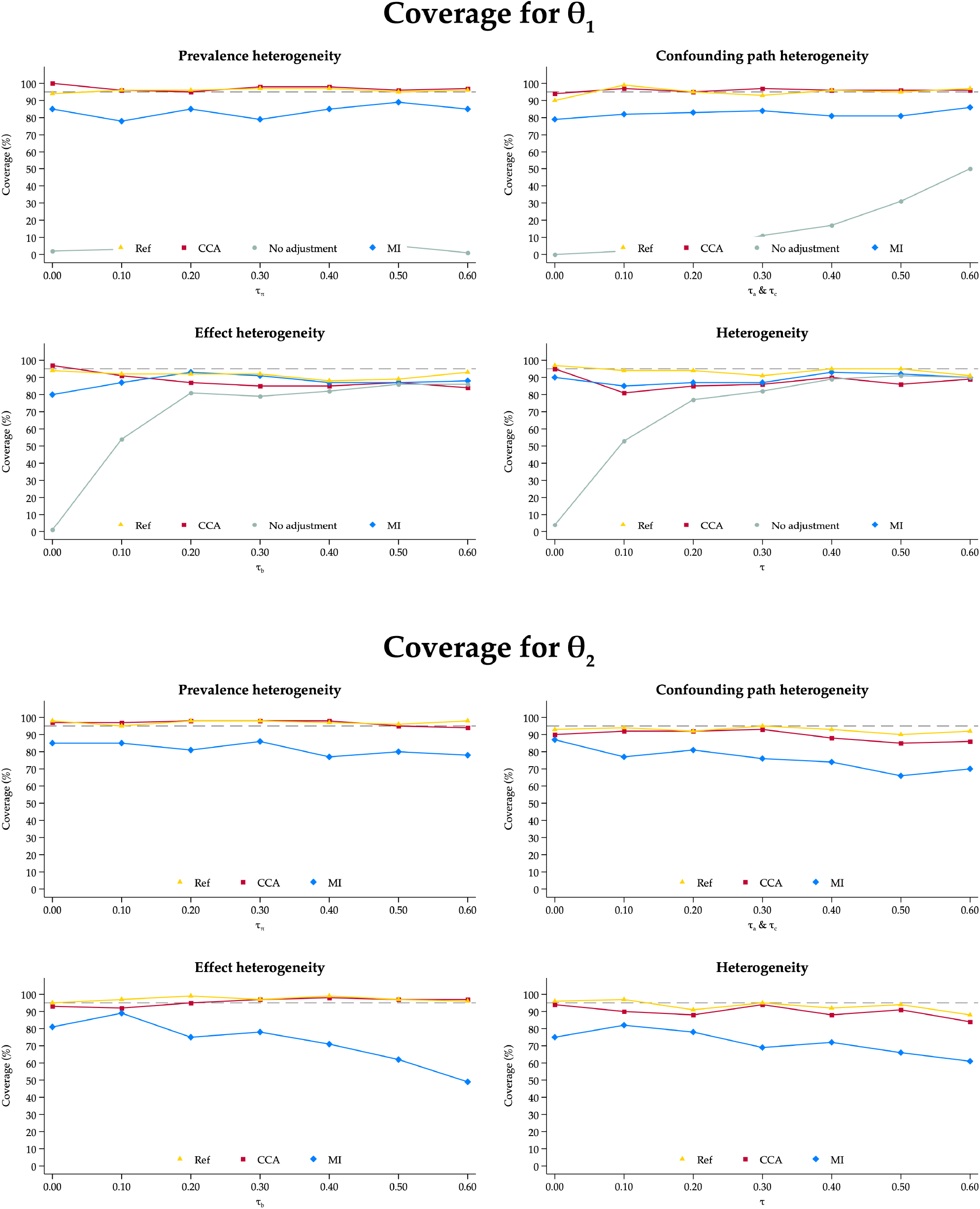
Coverage for *θ*_1_ and *θ*_2_

#### 4.3 Increased number of imputations

**Figure S8:**
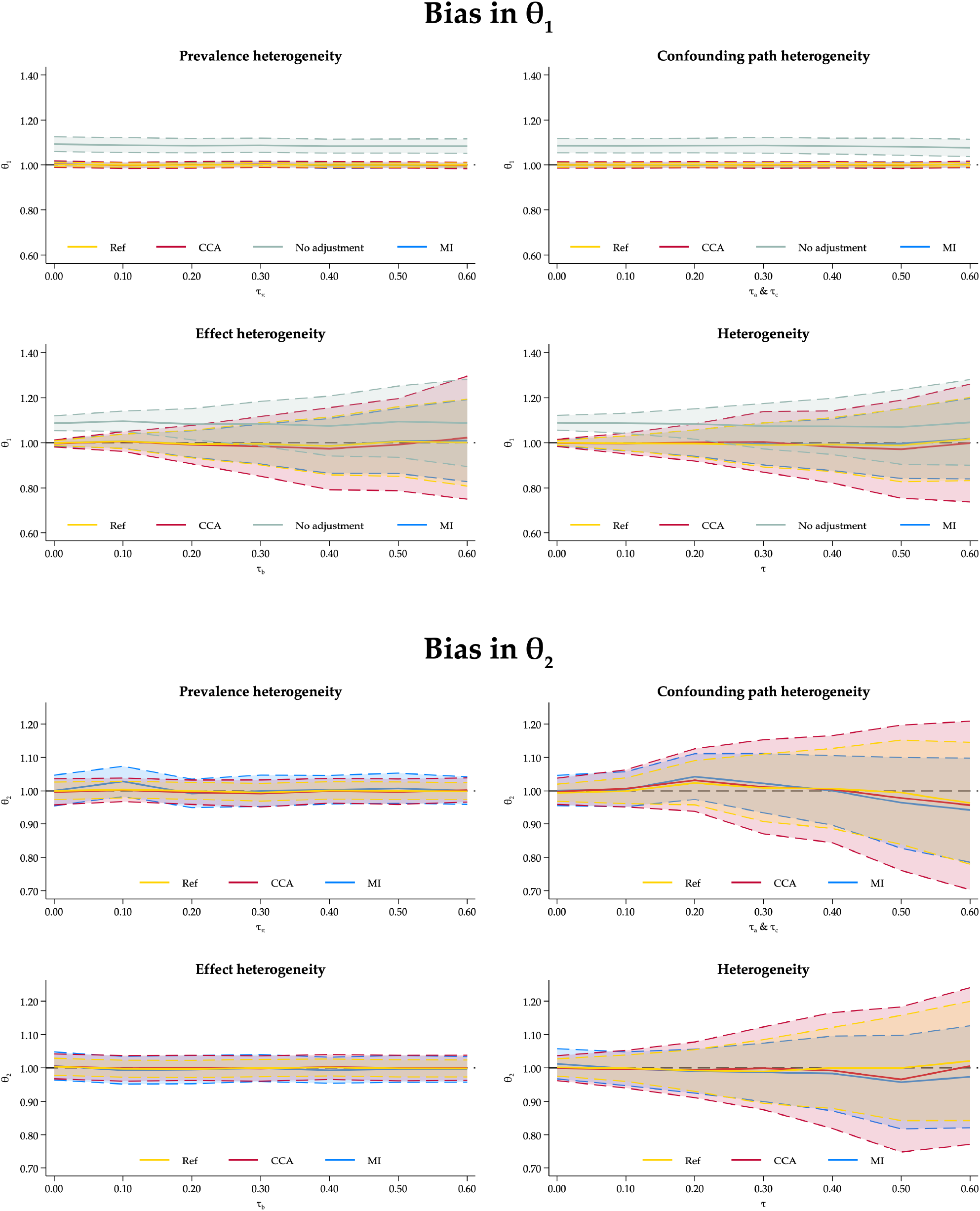
Bias for *θ*_1_ and *θ*_2_

**Figure S9:**
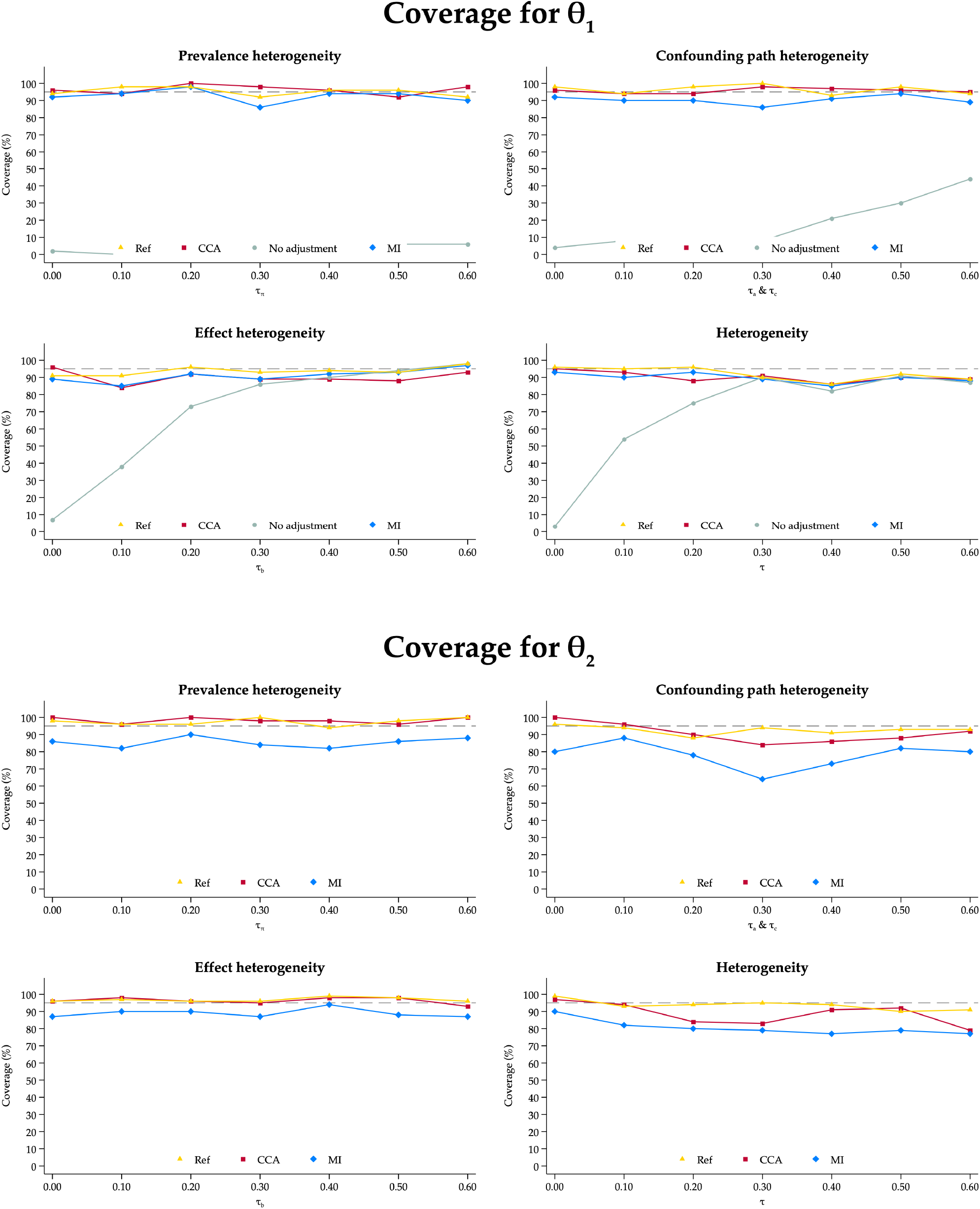
Coverage for *θ*_1_ and *θ*_2_

